# Accuracy of lung and abdominal ultrasound for tuberculosis diagnosis: a prospective cohort study from India

**DOI:** 10.1101/2024.06.07.24308357

**Authors:** Stefan Fabian Weber, Rebecca Wolf, Katharina Manten, Balamugesh Thangakunam, Barney Isaac, Deepa Shankar, Divya Mangal, Amit Kumar Dutta, Leena Robinson Vimala, Aparna Irodi, Frank Tobian, Lisa Köppel, Julia Selena Beck, Peter Wolf, Sabine Bélard, Claudia Maria Denkinger, Devasahayam Jesudas Christopher, And the ALL POCUS TB India study group, Mary Gaeddert, Lisa Ruby, Bharath Karthikeyan, Arin Natania, Sai Vijaysree, Sangeeth Priyadarshan Veluchamy Rathakrishnan

**Author notes:** corresponding: DJ Christopher, CMC Vellore, Ranipet Campus, Vellore 632004, India. shared (Claudia Maria Denkinger, Devasahayam Jesudas Christopher). Study group.

## Abstract

**Background:** Point-of care ultrasound is considered to hold promise in tuberculosis (TB) screening. However, most available abdominal ultrasound data focuses on HIV-infected cohorts and for lung ultrasound (LUS) data is very sparse. We aimed to determine accuracy of lung and abdominal ultrasound in a cohort of presumed TB in a tertiary care hospital in India.

**Methods:** Adult patients with presumed TB were enrolled prospectively and underwent a comprehensive ultrasound evaluation. Accuracy of individual and a predetermined combination of findings was determined against a TB reference standard (mycobacterial culture and PCR). Diagnostic potential of a multi-variable model combining clinical and ultrasound findings was explored using generalized mixed methods and random forest approach. (German trial registry DRKS00026636)

**Findings:** We included 541 participants of whom 102 (19%) had TB and 1% had HIV. “Focused assessment with sonography for HIV-associated tuberculosis” (FASH) showed moderate sensitivity (51%, 95%-CI 41-60) and specificity (70%, 95%-CI 66-74). Small consolidations on LUS showed high sensitivity (98%, 95%-CI 93-99), but were unspecific (14%, 95%-CI 11-18). Exploratory LUS variations showed higher specificity (e.g., large apical consolidations: sensitivity 22%, specificity 86%). Predictive modelling for ultrasound and clinical variables revealed an Area Under the Curve of 0.79 in the receiving operator curve.

**Interpretation:** Accuracy of ultrasound does not meet requirements of a stand-alone diagnostic or screening test. However, accuracy for some ultrasound findings is comparable with CXR. Additionally, ultrasound may aid disease severity assessment and microbiological sampling strategies. Research into alternative analyses (e.g., artificial intelligence) may enable wider applications.

**Funding:** Grant TTU 02.911, German Center for Infection Research (Deutsches Zentrum für Infektionsforschung).

**Research in context:** Before undertaking the study, LUS for TB had been assessed in a small number of studies limited with uncertain ultrasound characterization of TB-related findings with lack of adequate terminology and unclear specificity for TB. FASH-studies in HIV+ and few studies in HIV- have shown moderate sensitivity and specificity, but the study design and reference standards were not robust enough for generalizability.

Our ultrasound study of LUS and FASH has a prospective cohort from a TB-endemic setting (India), we recruited 541 participants, the largest such cohort. This study brings to the body of evidence novel findings, backed by a robust study design and using a comprehensive reference standard. We were able to describe accuracy in a predominantly HIV-negative cohort of patients with presumed TB disease and compared our index testing protocol with the CXR, the most commonly used imaging modality. The implications from our study were that no single finding or combination of findings on LUS reached target product profiles (TPP) proposed by WHO, suggesting LUS as triage or diagnostic tool is too non-specific. The FASH accuracy in our study was in line with previous data, this study provides for it a solid foundation. The accuracy of submodules of LUS and FASH reaches that of CXR. Roles for ultrasound in TB care may lie in aiding diagnosis, assessment of disease severity, guiding of microbiological sampling or therapy monitoring. Further studies should focus on the accuracy of LUS in people living with HIV and children, evaluating ultrasound as a part of a diagnostic algorithm and the use of artificial intelligence to improve the yield of TB-POCUS.

## Introduction

The challenge of tuberculosis (TB) diagnosis persists, as evidenced by the diagnostic gap of 26% (global TB incidence in 2022 10.6 million & notified cases 7.8 million). (1) Access to a diagnostic test is the prerequisite to receive treatment. In order to improve diagnosis, the World Health Organization (WHO) called for diagnostic tests to meet target product profiles (TPP), which for a TB triage test should meet a minimum sensitivity of 90% and specificity of 70%. (2) WHO recommended triage tests include chest x-ray (CXR). However, CXR, despite becoming more accessible due to innovation such as ultra-portable CXR systems (3) assisted by computer assisted detection (CAD) using artificial intelligence (AI) (4), is not widely available due to equipment and infrastructure costs.

Point-of-care ultrasound (POCUS), especially in HIV-related TB, has emerged as an addition to the diagnostic toolbox (5). “Focused assessment with sonography for HIV-associated tuberculosis” (FASH) is a protocol designed for HIV-related TB in resource constraint settings assessing for pericardial (>1cm) and pleural effusions (any size), hepato-splenic micro-abscesses and abdominal lymphadenopathy (6). A systematic review showed moderate accuracy, but studies were heterogeneous (7). However, data are scarce regarding its application in HIV-uninfected populations and the value of data is curtailed by suboptimal study design and the sensitivity ranged from 36-39% and specificity from 70-89%. (8-10) Lung ultrasound (LUS) has been a focus of TB-POCUS research recently and ultrasound findings, such as “subpleural nodules” and “miliary pattern” have been proposed as associated with pulmonary TB (PTB) (11, 12)). A systematic review summarized available studies suggesting a high sensitivity (e.g., subpleural nodules (SUN) up to 97%) but specificity remained unclear, mostly due to the lack of adequate control groups. (13) A further issue with current LUS research is comparability due to the varying definitions used and also entities such as SUNs lack a pathophysiological correlate.

Other possible targets for POCUS, such as lymphadenopathy in the internal mammary region, mediastinal lymph nodes (14-16), or peritoneal changes (17), have only been described in case studies or series. Their prevalence and potential value for POCUS applications remains to be elucidated in prospective studies with robust reference standards.

This study aims to investigate the accuracy of the FASH protocol using a rigorous study design in a representative study population within a referral hospital in a high TB burden setting. Additionally, we sought to explore the accuracy of LUS and assess other novel ultrasound targets.

## Methods

### Study design and participants

ALL POCUS TB (Abdominal and lung point-of-care ultrasound for tuberculosis) was a single-center, prospective diagnostic accuracy study conducted at an Indian referral hospital (Christian Medical College (CMC), Vellore). Participants were recruited from the Department of Pulmonary Medicine and referrals from the Department of Gastroenterology. Study staff reviewed charts of in- and out-patients to identify those meeting inclusion criteria. We recruited consecutive adults ≥18 years with presumed TB disease for whom a TB workup was initiated by attending physicians and who were positive in the WHO TB four-symptom screen (cough ≥2 weeks (any duration in HIV), fever, weight loss, night sweats (18)). We excluded patients who were already microbiologically confirmed at the time of screening, who had been started on anti-TB therapy (ATT) and those that had taken any TB-active medication within the previous 6 months. The study was approved by CMC Vellore and University Heidelberg institutional review boards (CMC IRB ID 14342; Heidelberg ID S-314/2021). The study was conducted according to Good Clinical Practice (GCP) guidelines and the Helsinki declaration. Informed consent was obtained from all participants.

The study was registered in the German trial registry DRKS (ID: DRKS00026636). This study conforms to the Standards for Reporting of Diagnostic Accuracy Studies (STARD) reporting guidelines, see STARD checklist in supplement. (19)

A parallel, collaborative study was also conducted in German referral hospitals. Methods were similar between countries but participant populations differed substantially, therefore the results from Germany are published separately.

### Procedures

A standardized questionnaire was used to collect demographic data, detailed symptom and medical history. Study laboratory tests included HIV-serology (positive if positive current or self-reported prior positive test), HbA1c (diabetes positive if HbA1c if ≥6.5% or self-reported prior diagnosis) and C-reactive protein (CRP).

For TB, at least two respiratory samples (two sputa or a sputum and a bronchoalveolar lavage (BAL) sample) were tested with liquid mycobacterial culture (BD BACTEC™ MGIT™ 960, pooling was possible) and at least one respiratory sample was tested via Xpert® MTB/RIF Ultra cartridges. One spot urine sample (>30ml, centrifuged) was tested via Xpert Ultra. Other investigations, invasive samples and imaging was performed at the discretion of the attending physician. Relevant hospital data and discharge information was collected from the hospital records to identify alternative diagnoses for participants considered not to have TB.

Repeat sputum testing was offered to participants who initially did not test positive for TB and who were not empirically treated for TB, but continued to have symptoms at follow-up at least two months after presentation.

### Index test

FASH: The pre-defined study POCUS protocol included FASH (6). In addition, we added pericardial and pleural effusion measurements (estimate adapted from Goecke et. al. (20)) to assess if lower cut-off for pericardial effusion or an added cut-off for pleural effusion affects accuracy (FASH definition and variations, see Table 1).

LUS: For LUS we adapted existing lung protocols for 14 lung zones (Figure 1) (21). All zones were scanned both vertical and parallel to the intercostal spaces (ICS) with a linear probe (if at least two A-lines were visualized, otherwise we switched to curved probe). We assessed each zone for the presence of A-lines, B-lines, pleural irregularities, pleural effusions, subpleural consolidations (SPCs) and miliary pattern. In addition to these pre-defined findings, we recorded the number, size and location of findings for an exploratory analysis of accuracy, see Table 1 for definitions.

**1.**
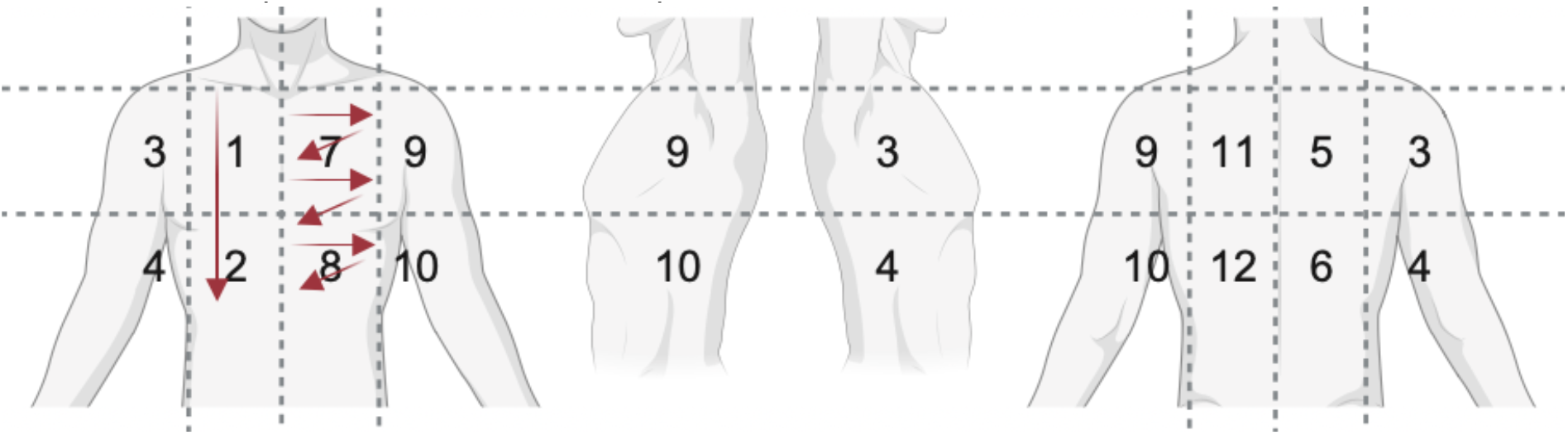
schematic of lung zones and probe movement. Zone 1/2: example for longitudinal sweep, zone 7/8: example for intercostal sweeps

**2.**
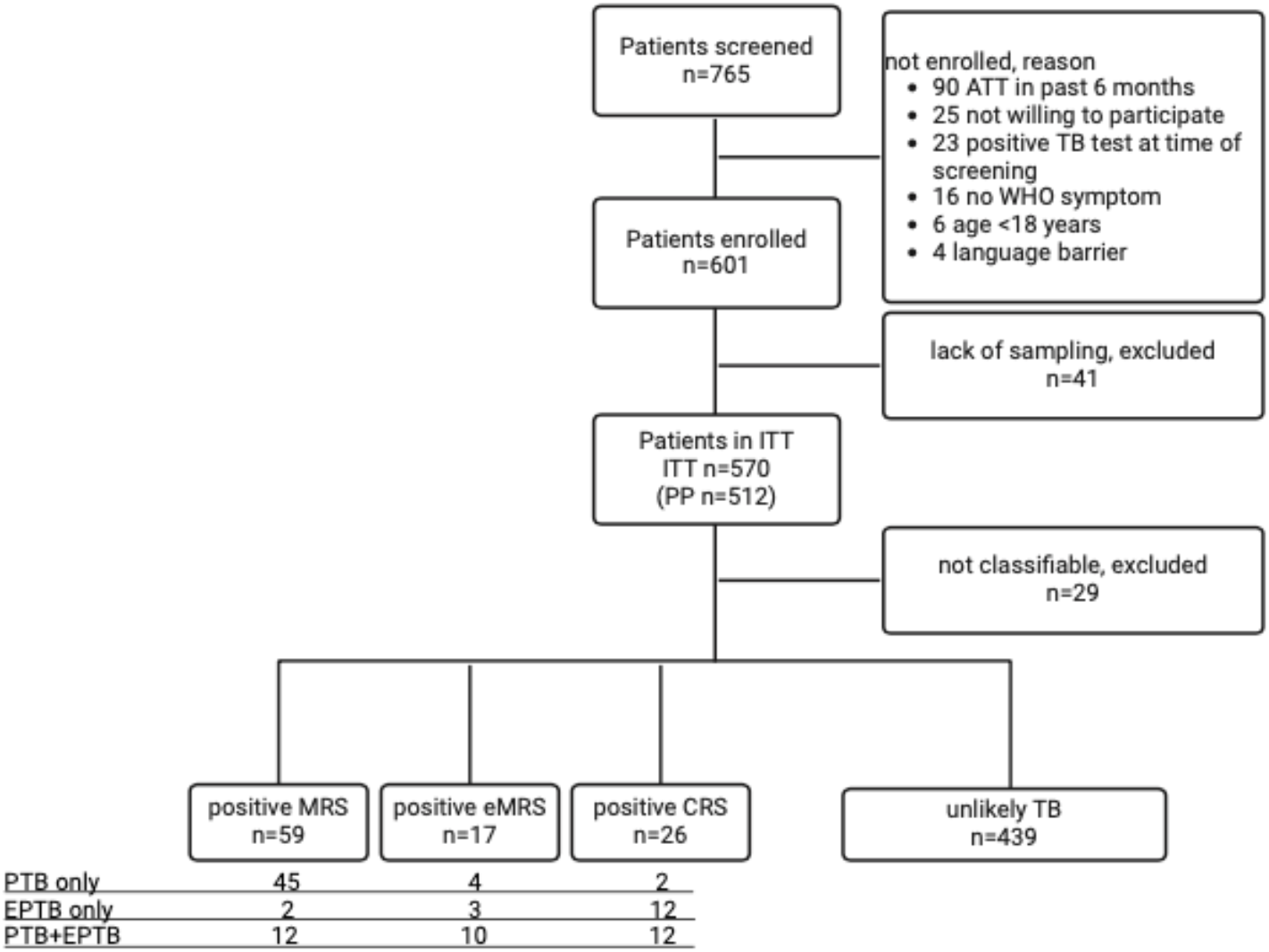
Flow chart for study recruitment and reference standard categories ATT anti-tuberculous treatment; WHO symptom cough >2 weeks, fever, night sweats, weight loss; PP per protocol cohort; ITT intention-to-test cohort; MRS microbiological reference standard; eMRS extended MRS; CRS composite reference standard

We assessed for IMNs in the parasternal ICSs, peritoneal or omental thickening in all abdominal quadrants and in the trans-costal liver view; in the right lower quadrant we assessed for ileocecal thickening > 4mm. Using a phased array probe, we attempted to visualize mediastinal structures from the suprasternal notch and left parasternal view for lymphadenopathy. In participants with clinically presumed peripheral TB-lymphadenopathy, we scanned respective areas. For details see Table 1.

We used an Edge II (Fujifilm Sonosite, Bothel, United States of America) with rC60xi (curved), L38xi (linear) and rP19x (phased) probes. Ultrasound clips were recorded for all standard views. Ultrasound was performed on the day of enrolment by non-radiologist clinicians (RW, KM) trained in the ultrasound procedures by SFW. Proficiency was ensured by trial clips evaluated for protocol adherence. Ultrasound investigators were blinded to participants’ reference standard test and imaging data. If an ultrasound finding was not adequately visualized, it was marked non-evaluable. To assess inter-rater agreement, a random sample of 15% of recorded ultrasound studies from all sites were evaluated by an additional rater independently (SFW, RW, KM) regarding the presence of SPC_≥1cm_, SPC_<1cm_, and FASH items.

### Comparator test

Available chest x-rays (recruitment ±2 weeks), were assessed by two senior Indian radiologists blinded to clinical and index test data (consensus read). Radiologists interpreted images as “CXR suggesting likely TB” or “CXR suggesting possible TB”, “suggestive of post-TB sequelae” (with or without active TB) or “not suggestive of active TB or post-TB sequelae” guided by published suggestions (22).

CRP as a predictor was evaluated at a cut-off of 5mg/l. (18)

### Reference standard and case definitions

Participants with complete sample set (i.e., two respiratory samples with TB-culture (pooled or separate) and TB-PCR as well as urine TB-PCR) were assigned to the per protocol (PP) population. Participants with at least one respiratory sample with TB-culture or two respiratory samples without culture, but with TB-PCR and also participants with only non-respiratory sample in presumed exclusive extra-pulmonary TB were assigned to the intention-to-test (ITT) population. All participants not fulfilling these criteria were excluded from further analyses.

We compared cohort characteristics between PP and ITT (available in the Supplement Table 1). A generalized linear mixed model approach with belonging to PP as binary covariate and a random effect accounting for individual variability revealed that belonging to PP or not does not statistically significantly affect TB diagnosis (p = 0.987). As no relevant differences in index test accuracy for FASH and SPC was observed between PP and ITT group, we report on the larger ITT cohort below. PP results are separately available in Supplement Table 5.

The reference standards for tuberculosis disease were defined as a microbiological reference standard (MRS), an extended MRS (eMRS) or a composite reference standard (CRS). TB was diagnosed if 1) the MRS was positive; i.e. any respiratory or urine sample was positive for *Mycobacterium tuberculosis* (MTB) by culture or PCR; or 2) the eMRS was positive, i.e. any other sample (e.g., biopsy) was positive for MTB on culture or PCR; or lastly 3) the not pre-defined CRS was positive, which meant that empirical ATT was started on clinical grounds (e.g., radiology, histopathology) and clinical improvement was documented under ATT. Trace positive PCR results were repeated and only considered positive if the second sample was also positive (with any semi-quantitative result).

Participants were considered unlikely to have TB if MRS, eMRS and CRS were negative AND at least one of the following criteria applied: follow-up with negative sputum, or symptom resolution, or a plausible other diagnosis (as per pre-defined list, see SAP in supplement). Remaining cases not falling into these categories underwent an outcome committee assessment (2 senior Indian (DJC, BT) and 2 senior German (CMD, PW) TB-physicians). The committee had access to clinical and diagnostic data but were blinded to the index test (POCUS). Ruling by consensus was either positive CRS, unlikely TB or unclassifiable (excluded from further analyses).

#### Statistics

A target sample size of 577 was calculated (inputs: sensitivity 60%, specificity 85%, prevalence 20%, precision 10%, dropout 20%). (23) The primary outcome for POCUS was sensitivity and specificity (including 95%-confidence intervals (95%-CI)) of the FASH protocol and individual LUS findings. We used CRS as the primary and most sensitive reference standard. In the analysis, we will refer to these as “TB cases”. Exploratory outcomes included an analysis of POCUS findings either individually or combined as well as subgroup analyses by diabetes or HIV status.

Descriptive data are presented as numbers and proportions (%) or median and interquartile range (IQR) as applicable. Significance testing was performed where applicable using X^2^-Test, Fisher’s exact test or Mann-Whitney U-test where applicable with significance level defined as p≤0·05. For rater agreement. To explore predictive performance of the data in an HIV-negative cohort, we applied subset selection and machine learning algorithms. For this purpose, we first excluded all HIV+ participants and performed reduction of dependency structures within the predictors using factor analysis. Through assessing and excluding specific correlating variables carrying similar information from all variables excluding CXR, we obtained a subset of the ultrasound and clinical variables. We then determined statistically significant predictors for TB status by means of Lasso Regression to punish less impactful predictors. To account for correlations within the predictors, we applied a random forest approach with five-fold cross validation to produce two prediction metrics, AUC (area under the curve) and ROC (Receiver Operating Characteristic) curve. Performance metrices were compared on the subsets of predictors following factor analysis and Lasso regression. Sensitivity analysis was also conducted, including CXR as additional predictor for both subsets.

We used Cohen’s kappa and a generalized linear mixed model to account for the different rater combinations and random error due to repeated measurements of individuals for the probability of agreement. Statistical analyses were performed using R with packages (openxlsx, REDCapR, ggplot2, dplyr, Hmisc) and Python (Version 3.6) with the libraries factor_analyzer, statsmodels and sklearn. We used RedCap (Version R 4.2.2 (24)) for data collection. Figures were created using biorender.com and R packages.

## Results

Between 18^th^ April 2022 and 29^th^ July 2023, we screened 765 patients. Of 601 enrolled participants, 512 (85%) were eligible for PP and 570 (95%) for the ITT analysis, 31/601 (5%) were excluded due to insufficient samples for PP or ITT allocation and another 29/601 (5%) were unclassifiable as per reference standard and were also excluded from further analyses. Here, we report the results on 541/601 (90%) participants in the ITT cohort.

The median age was 48 years, and 357/541 (66%) were male. Diabetes was present in 121/512 (24%) and HIV in 6/523 (1%). History of prior TB was reported by 99/540 (18%) and a current TB contact was reported in 5/535 (1%). Smoking was reported in 110/541 (20%). Considering the WHO 4-symptom screen, 483/541 (89%) reported cough, 48/541 (9%) night sweats, 230/541 (43%) fever and 292/541 (54%) weight loss.

In total, 102/541 (19%) of participants were diagnosed with TB (59 per MRS, additionally 17 per eMRS and additionally 26 per CRS), 439/541 (81%) were deemed unlikely to have TB. TB was limited to the lungs (PTB only) in 59/102 (58%) cases, limited to EPTB (no PTB) in 17/102 (17%) and concurrent EPTB with PTB was present in 26/102 (25%). Most frequent alternative other diagnoses were non-TB lung infections (102/439, 23%), asthma (92/439, 21%), chronic obstructive pulmonary disease (or other obstructive lung disease, 71/439, 16%) and post-TB sequelae (51/439, 12%), see Table 5.

All baseline characteristics of the study cohort for all participants, TB and unlikely TB cases based on CRS are provided in Table 2, data for MRS and eMRS are available in Supplement Table 2.

### Index Test

FASH_original_ had a sensitivity of 51% (95%-CI: 41-60) and specificity of 70% (95%-CI 66-74). Sensitivity was mostly driven by the most common FASH-component pleural effusion (172/541 (32%) of all participants, 47/102 (46%) of the TB cases and 125/439 (28%) of those unlikely to have TB). Other FASH-components were rare: abdominal lymph nodes were present in 15/541 (3%), pericardial effusions in 8/541 (1%), hypoechoic spleen lesions in 7/541 (1%, cf. Figure 3, g) and hypoechoic liver lesions in 5/541 (1%).

**3.**
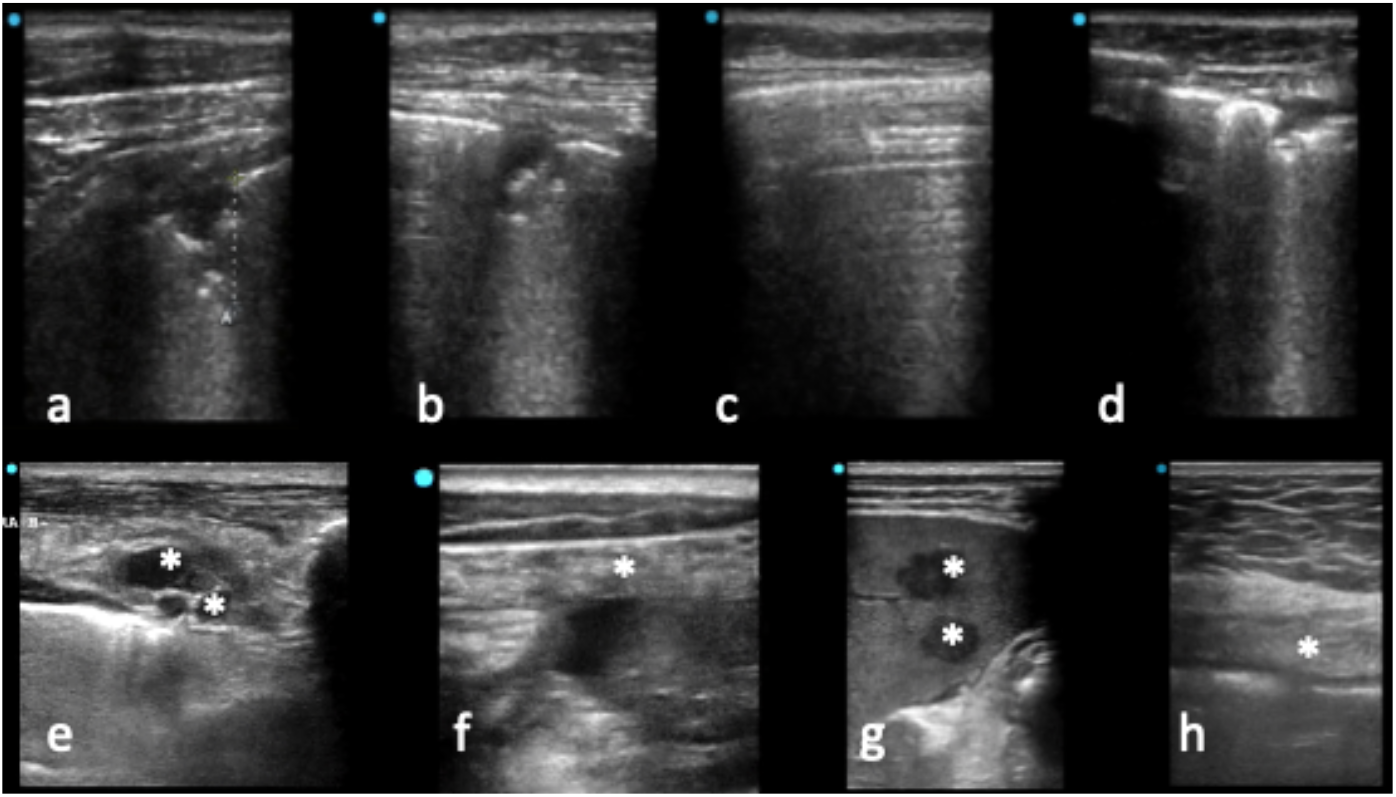
Panel: POCUS examples a+b: male in his 60s, DM+, HIV-, MRS+, SPC > and < 1cm c: male in his 20s, MRS+, normal lung area with A-lines neighboring consolidated area d: male in his 70s, DM-, HIV-, post-TB, now pseudomonal pneumonia with SPS <1cm e: male in his 50s, HIV-, DM+, CRS+, disseminated TB (pericarditis, pleural, peritoneal) here: internal mammary lymph nodes (asterisk) f+g: male in his 10s, HIV-, under anti-TNFalpha (spondyloarthropathy), MRS+, disseminated TB with omental thickening with hypoechoic lesions (asterisk, F); >10 hypoechoic spleen lesions 0·5-1·3cm (asterisk, G) h: male in his 60s, DM+, HIV-, CRS+, omental thickening with hypo-hyper-hypo-echoic layering (asterisk)

When exploring adaptations to FASH such as a minimum 600ml threshold to pleural effusions, FASH_pleural600ml_ sensitivity decreased to 35% (95%-CI: 27-45) but specificity increased to 84% (95%-CI: 81-87), which is also reflected in the larger estimated pleural effusion volume in participants with TB (TB cases median 914ml vs. unlikely TB: 462ml). Including ascites (FASH_ascites_) or reducing the pericardial effusion cut-off to 4mm (FASH_pericardium_) did not have a relevant effect on accuracy. Data for FASH and single FASH-components are provided in Table 3 and performance of additional variations are available in supplement Table 3. In positive FASH_original_ but unlikely TB, alternative diagnosis was lung neoplasia in 31/130 (24%) and non-TB lung infections 28/130 (22%) cases.

On LUS, SPCs_<1cm_ were very common in both participants with TB and those unlikely to have TB: (sensitivity 93%, specificity16%). In comparison, larger SPCs_≥1cm_ were less common in TB cases (sensitivity 72%) and also in unlikely TB (specificity 55%). When exploring the location or number of zones affected by SPCs, we found higher specificity, but lower sensitivity if only considering SPCs_≥1cm_ in the apical lung zones (sensitivity 22%, specificity 86%) or SPCs_≥1cm_ in at least three lung zones (sensitivity 38%, specificity 79%).

Miliary pattern was seen with similar frequency in both groups: 13/102 (13%) in TB cases and 50/439 (11%) in unlikely TB. B-lines were present in 53/102 (52%) of TB cases and in 193/439 (44%) of unlikely TB. Data for LUS findings and variations of size, number and location of findings is shown in Table 3, further details are available in Supplement Table 3.

The most common differential diagnoses for SPCs_<1cm_ were non-TB lung infections (96/369, 26%), asthma (71/369, 19%) and COPD (67/369, 18%). For SPCs_≥1cm_, non-TB lung infections (67/209, 32%) and post-TB sequelae (38/209, 18%) were the most common diagnoses. A miliary pattern was mostly seen in participants with non-TB lung infection and ILD (15/50, 30% each), see Table 5.

When considering other targets, IMNs (cf. Figure 3, e), pleural thickening, peritoneal thickening and intestinal thickening in the right lower quadrant were all present in <20% of participants but more common in TB cases: prevalence of IMNs in TB cases was 16/102 (16%) vs. in unlikely TB 18/439 (4%); prevalence of pleural thickening was 14/102 (14%) in TB cases vs. 30/439 (7%) in unlikely TB; peritoneal thickening prevalence in TB cases was 11/102 (11%) vs. unlikely TB with 3/439 (1%); c.f., Figure 3, f, h; prevalence of intestinal thickening in the lower right quadrant 9/102 (9%) in TB cases and 6/438 (1%) in unlikely TB.

### Subgroup analyses by HIV and diabetes status

Ultrasound in diabetes: pleural effusions were slightly more common in participants with diabetes (36%) than in those without (31%) with otherwise no relevant differences between groups regarding FASH findings or SPCs_<1cm_ and SPCs_≥1cm_. All subgroup analyses for ultrasound stratified by diabetes status are provided in Table 4. For ultrasound in HIV+, case numbers were insufficient for analysis (n=7), HIV-stratified data is available in Supplement Table 4.

Interobserver agreement calculations for ultrasound view rating (see methods) yielded an overall Cohen’s kappa of 0·71, from the generalized linear mixed we inferred the probability of agreement to be 98·8%, which strongly supports coherence in rater decisions.

### Comparator tests

“CXR suggesting likely TB” had a sensitivity of 34% (95%-CI 25-44) and a specificity of 89% (95%-CI 86-92)), comparable with FASH_peural600ml_ which at 35% sensitivity was 84% specific. “CXR suggesting possible TB” (includes “CXR suggesting likely TB”) had a sensitivity of 81% (95%-CI 72-87) and specificity of 58% (95%-CI 53-62)), which was in the range of any SPCs_≥1cm_, which at 72% sensitivity was 55% specific. In comparison, CRP had a sensitivity of 82% (95%-CI 73-89) and a specificity of 53% (95%-CI 48-58). Correlation and Venn diagrams for selected ultrasound findings vs. CXR and CRP are provided in Figure 4.

**4.**
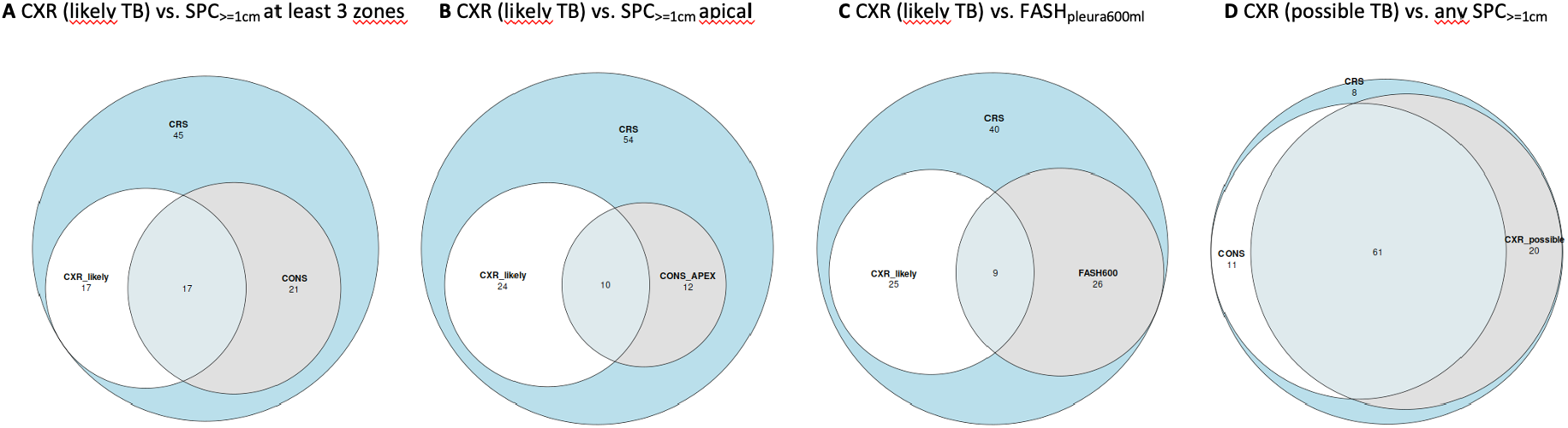
Venn diagrams of overlap between lung ultrasound findings and chest x-ray Color code: White area: unlikely TB Light gray circle (left): ultrasound positive Dark gray circle (right): chest x-ray positive Overlap gray circles: ultrasound and chest x-ray positive CXR: chest x-ray FASH_pleural600ml_: Focused assessment with sonography for HIV-associated tuberculosis with a pleural effusion threshold of 600ml

### Predictive modeling analysis

Including all variables (except CXR) derived after the factor analysis reached an AUC of 0·79. Lasso regression led to a further reduction from 22 predictors to 10 significant variables (see supplement section “Predictive modelling analysis”), while increasing the AUC to 0·75 only. Combining CXR with all variables after factor analysis achieved an AUC of 0·82. Using CXR only the model reached an AUC of 0·7. Comparing ROC curves, CXR revealed a slightly better performance specifically in the high sensitivity aspects of the ROC curve. ROC curves are provided in Figure 5, further details and all variables used in the model are provided in Supplement.

**5.**
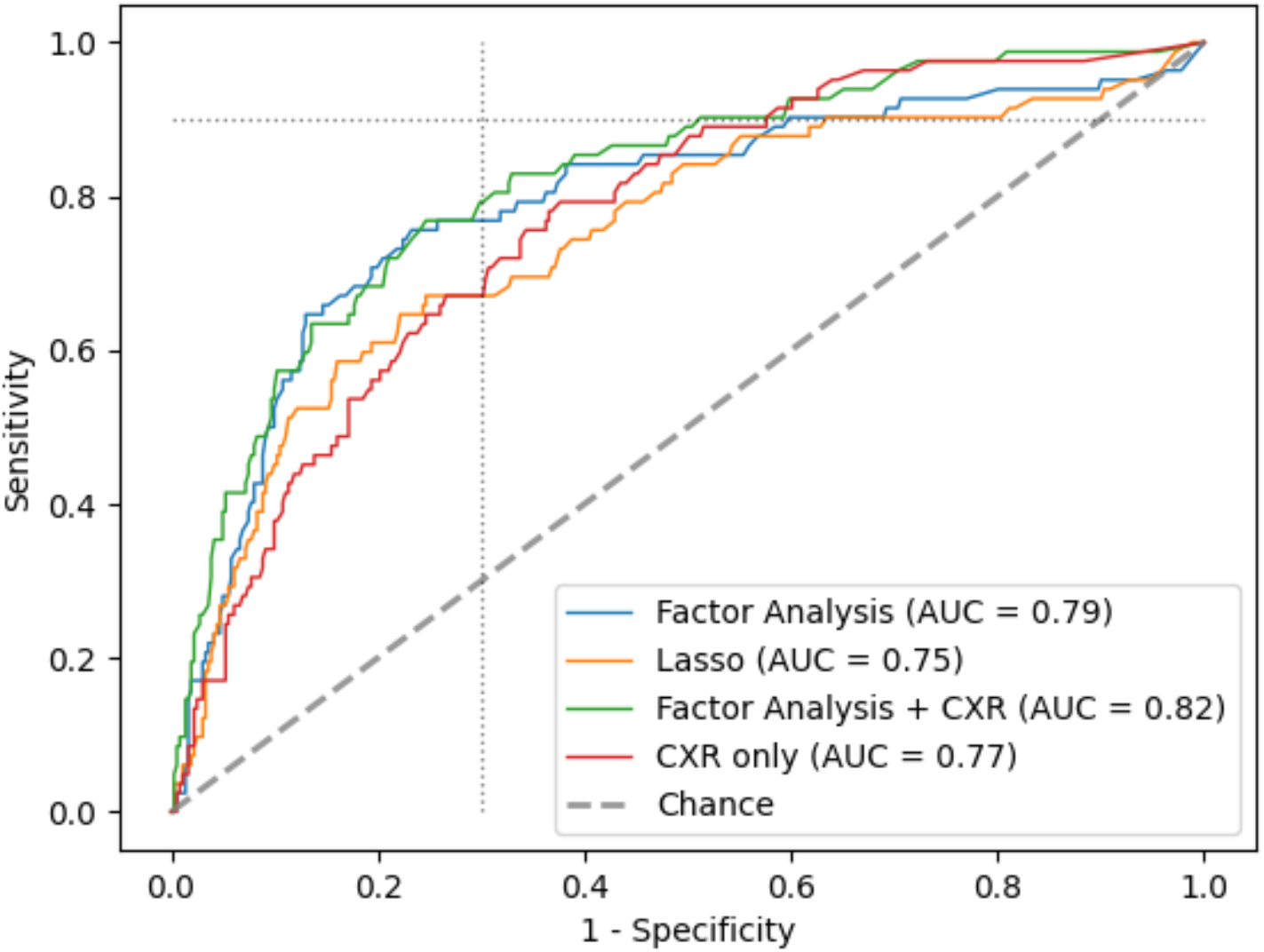
Receiving operator curve for ultrasound, CXR and other participant variables for TB diagnosis Dotted lines: cut-off for 70% specificity and 90% sensitivity AUC: area under the curve CXR: chest x-ray Variables in factor analysis and Lasso analysis, see Supplement

No adverse events were observed as part of the reference standard or index test.

## Discussion

This multi-center study reports the largest cohort investigated for LUS, FASH and other thoracic and abdominal findings in presumed TB prospectively.

With regards to FASH in our predominantly HIV-uninfected cohort, our data showed moderate sensitivity (51%) and specificity (70%), which is more sensitive and similarly specific as compared to a previous Indian study (39% sensitive, 70% specific (10)) and more sensitive but less specific than a previous study from South Africa (36% sensitive, 89% specific (9)), both of which have limitations with regards to unrepresentative control groups. A systematic review on ultrasound for EPTB including participants irrespective of HIV-status, found a pooled sensitivity of 72% and specificity of 77%. (25) In our cohort, sensitivity of FASH was mostly driven by larger pleural effusions, which contrasts to FASH in HIV-infected individuals, where abdominal lymphadenopathy (range 36-86%) and spleen lesions (range 13-62%) were more common in a Cochrane review. (7) Other EPTB findings were rare suggesting that they may not be relevant in a POCUS use case in HIV-negative individuals. Introducing a cut-off for pleural effusions appears to increase specificity by reducing detection of minor effusions: accuracy of FASH_pleural600ml_ was comparable with CXR, when an interpretation of “CXR suggesting likely TB” was chosen. However, a possible added value of FASH may be to detect different TB-cases than those detected by CXR (25% of TB-cases would have been detected by FASH_pleural600ml_ but not by “CXR suggesting likely TB” (Figure 4, A). Additional value of FASH in HIV-uninfected individuals may be an exploration of disease spread or targeting diagnostic interventions.

LUS had variable accuracy depending on the size, location and number of findings investigated. SPC_≥1cm_ in the apical lung zones (22% sensitive, 86% specific) or SPC_≥1cm_ in at least three lung zones (38% sensitive, 79% specific) were in a similar accuracy range as “CXR suggesting likely TB” (sensitivity 34%, specificity 89%). Any SPC_≥1cm_ (sensitivity 72%, specificity 55%) was slightly less sensitive but comparably specific as “CXR suggesting possible TB” (81% sensitive, 58% specific). Correlation of LUS vs. CXR was highest for any SPC_≥1cm_ vs. “CXR suggesting possible TB”, see Figure 4, D.

In a systematic screening setting, CXR has shown superior accuracy to our CXR interpretation (pooled sensitivity of 85% and specificity of 96% (26)), however data on CXR in triage after symptom screening is limited but comparable accuracy as our study CXR interpretation (90% sensitive, 56% specific (27)). LUS findings of higher specificity may justify possible use of LUS in certain settings in a diagnostic algorithm (e.g., forwarding LUS-positive cases for confirmatory testing and performing additional screening tests if negative), but overall no finding and no variation in size, location of number of zones affected surpassed WHO TPP goals (2) as a stand-alone test. Additional uses of LUS may be the assessment of disease severity.

Low specificity in our cohort for LUS and EPTB findings were caused by other lung infections, post-TB sequelae, lung neoplasia and obstructive lung disorders, e.g., due to bronchiectasis (cf., Table 5), which were showing similar ultrasound appearance as tuberculosis. Awareness of these differential diagnoses is warranted, especially if TB cannot be microbiologically confirmed.

### Other ultrasound findings

SPCs_<1cm_ with oval and regular shape, hypoechoic echogenicity and posterior enhancement (i.e., the characteristics suggested previously as subpleural nodules (11, 12)) were seen only in 12/541 (2%) of cases and irregular, non-oval shape resembling “mini shreds” were revealed by linear ultrasound in most cases (cf. Figure 3, a, b). These were unspecific in our cohort and likely represent lung tissue changes rather than solid nodules and we suggest not to use the term subpleural nodule without further evidence for a separate pathophysiologic entity.

Other findings like IMNs, pleural or peritoneal thickening were rare, but if found may guide clinical decision making and sampling strategies in difficult-to-diagnose cases. Unlike in children (16), mediastinal lymph nodes were not reliably assessable via suprasternal or parasternal sonography.

### Predictive Modeling analysis

This work aimed at exploring the ability to utilize ultrasound in combination with clinical variables for TB prediction from the collected dataset. The modeling exploration for prediction performed on this dataset confirmed the only moderate performance of ultrasound obtained from data exploration earlier. Else, the difference in ROC curves was marginal and may be attributable to randomness, suggesting comparable performance between ultrasound and CXR. For developing a diagnostic algorithm for possible clinical implementation, further work accounting for the inherent correlation structure of predictors, as well as the variety of extrapulmonary TB, as well as external validation with a separate study dataset is needed.

### Limitations and strengths

Limitations of our study was that CMC Vellore is a tertiary (referral) center, and the cohort with often advanced disease may not be fully representative of peripheral settings.

Strengths of our study was the large sample size, the robust reference standard (over 90% with at least two sputa, 75% within MRS or eMRS) and follow-up to optimize categorization by case definition. Case allocation was also supported by an expert committee, reducing number of cases where TB could not be ruled out to a minimum. The index test was performed in a comprehensive way and video documentation was performed for all normal and abnormal findings. Standardization of the protocol and reproducibility of ultrasound interpretation was confirmed by a high inter-observer agreement. For CXR reading, blinded review by TB-experienced radiologists enabled a representative comparison between POCUS and CXR. These strengths lead to a high level of generalizability of our data.

## Conclusions

Sub-components of our study ultrasound protocol showed comparable accuracy to CXR. While CXR itself is an imperfect diagnostic means for TB diagnosis, it has the disadvantage of low availability in peripheral settings and also uses ionizing radiation, whereas ultrasound has higher availability than CXR. Overall, however, accuracy of LUS and FASH in HIV-uninfected individuals does not achieve diagnostic performance required of a stand-alone diagnostic tool for TB and it does not facilitate a clear rule-in or rule-out of TB. Algorithmic approaches may provide opportunities to implement POCUS in diagnostic pathways and possible additional use cases may be delineation of disease extent, guiding sampling strategies and monitoring under anti-TB treatment, especially if no other monitoring modalities are available (this aspect will be addressed in future analyses of our longitudinal dataset). Remaining gaps in TB-POCUS are the lack of solid data for FASH and LUS in HIV-infected individuals as well as LUS in younger populations (children, adolescents), where pre-existing lung damage (e.g., due to smoking, dust inhalation, past PTB etc.) lowering specificity of LUS may be less prevalent.

## Supporting information

Tables for Manuscript

## Data Availability

Deidentified data will be made available upon reasonable request and provided in accordance to corresponding regulatory requirements.

## Acknowledgements

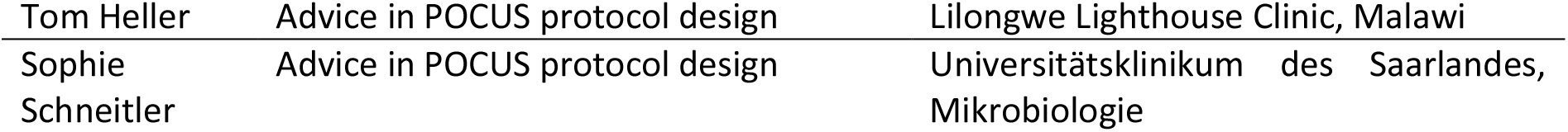

## Authors’ contributions

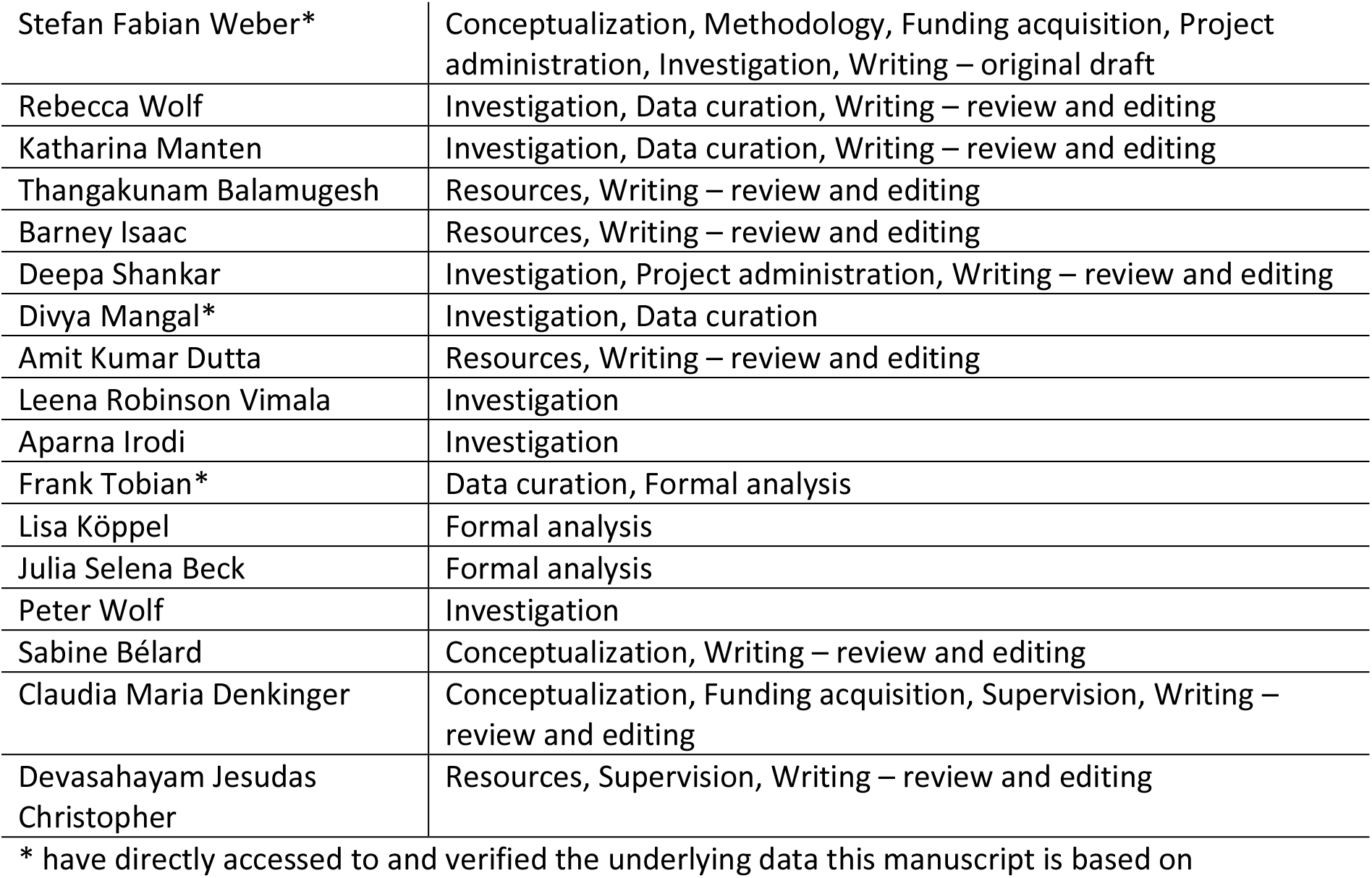

**Tables: see separate word file**

**Figures**

## Supplement

<u>Study protocol: DOI</u>

<u>SAP: DOI</u>

<u>Supplement Table 1 comparison of PP vs. ITT</u>

<u>Supplement Table 2 extended (ITT)</u>

<u>Supplement Table 3 extended (ITT)</u>

<u>Supplement Table 4 - POCUS – stratified by HIV and diabetes</u>

<u>Supplement Table 5 PP data</u>

<u>Interrater analysis</u>

<u>Predictive modelling analysis</u>

