## Supplementary material for "Accuracy of lung and abdominal ultrasound for tuberculosis diagnosis: a prospective cohort study from India": Tables for Manuscript

**Tables for the Manuscript**

| **Table 1 - ultrasound findings definitions and FASH variations** | | | |
| --- | --- | --- | --- |
|  | Definition | Further characteristics assessed | Location |
| SPC | Subpleural consolidations, hypoechoic or mixed-echoic lesions originating from the visceral pleura | maximum vertical dimension or translobar if not measurable, bronchograms | 14 lung zones |
| B-lines | Vertical lines originating from visceral pleura and extending to >50% of viewfield, minimum of 3 per lung zone | NA | 14 lung zones |
| Miliary pattern | B-lines, comet artefacts, pleural irregularities, small subpleural consolidations affecting all lung fields | NA | 14 lung zones |
| Pleural thickening | Thickening of parietal or peritoneal pleura | Laminar thickening or nodular thickening, maximum thickness | 14 lung zones |
| Pleural fluid | Content of hypoechoic or mixed echoic echogenicity between parietal and visceral pleura | Echogenicity, measurement of basal lung to diaphragm distance and maximum cranio-caudal distance, estimation of volume (Goecke et al., 1990) | Bilateral pleural recessus |
| Peritoneal thickening | Thickening of the peritoneal lining in the parietal, visceral or omental layers | Maximum thickness, inclusion of hypoechoic lesions, laminar or nodular thickening | 4 abdominal quadrants, transcostal liver view |
| Intestinal thickening | Thickening of the intestinal wall in the terminal ileum region exceeding 4mm | - | Right lower abdominal quadrant |
| Peritoneal fluid | Free abdominal fluid | Amount (small, moderate, large), echogenicity, content | Morrison, Koller and retrovesical pouch as well as inter-intestinal |
| Pericardial fluid | Pericardial fluid ≥4mm | Measurement, echogenicity, content | Sub-xiphoidal view |
| Spleen lesions | Intra-parenchymal spleen lesions   - Hypoechoic <1·5cm - Hypoechoic ≥1·5cm - echogenic | Measurement, count | Left flank |
| Liver lesions | Intra-parenchymal hypoechoic lesions | Measurement, count | Subcostal, transcostal |
| Abdominal lymph nodes | Lymph nodes ≥1·5cm any dimension | Measurement, architecture, bulking | Liver and spleen hilum, peri-pancreatic, para-aortic |
| Internal mammary lymph nodes | Lymph nodes ≥0·5cm, any dimension | Measurement, side, architecture, bulking | Parasternal intercostal spaces |
| Mediastinal lymph nodes | Lymph nodes ≥1·5cm, any dimension | Measurement, side, architecture, bulking | Suprasternal view, parasternal view |
| FASH_original_ | - abdominal lymph nodes ≥1·5cm - hypoechoic liver or spleen lesions - pleural effusion, any amount - pericardial effusion ≥1cm | | |
| FASH_ascites_ | - abdominal lymph nodes ≥1·5cm - hypoechoic liver or spleen lesions - pleural effusion, any amount - pericardial effusion ≥1cm - ascites, any amount | | |
| FASH_pericardium_ | - abdominal lymph nodes ≥1·5cm - hypoechoic liver or spleen lesions - pleural effusion, any amount - pericardial effusion ≥0·4cm | | |
| FASH_pleural_ | - abdominal lymph nodes ≥1·5cm - hypoechoic liver or spleen lesions - pleural effusion, minimum amount specified - pericardial effusion ≥1cm | | |
| **Legend**  SPC, subpleural consolidations;  FASH, focused assessment with sonography for HIV-associated tuberculosis  NA, not applicable | | | |

| **Table 2 - patient characteristics and reference standard testing** | | | | |
| --- | --- | --- | --- | --- |
| Variables in n, median, IQR, (%) | | All participants (n=541) | CRS positive (n=102) | Unlikely TB (n=439) |
| Age in years | | 48 [35;59] (N=541) | 44 [29;56] (N=102) | 49 [36;60] (N=439) |
| Sex male | | 357/541 (66%) | 65/102 (64%) | 292/439 (67%) |
| Body mass index (kg/m^2) | | 21·6 [18·4;24·9] (N=541) | 19·4 [17·1;23·1] (N=102) | 22 [19·1;25·2] (N=439) |
| Final diabetes status | | 121/512 (24%) | 23/94 (24%) | 98/418 (23%) |
| Final HIV status | | 6/523 (1%) | 1/101 (1%) | 5/422 (1%) |
| History of previous TB disease | | 99/540 (18%) | 11/102 (11%) | 88/438 (20%) |
| History of current TB-contact | | 5/535 (1%) | 3/102 (3%) | 2/433 (0%) |
| Symptoms | | | | |
| Cough | | 483/541 (89%) | 89/102 (87%) | 394/439 (90%) |
| Hemoptysis | | 134/541 (25%) | 18/102 (18%) | 116/439 (26%) |
| Night sweats | | 48/541 (9%) | 16/102 (16%) | 32/439 (7%) |
| Fever | | 230/541 (43%) | 68/102 (67%) | 162/439 (37%) |
| Weight loss | | 292/541 (54%) | 74/102 (73%) | 218/439 (50%) |
| Fatigue | | 348/541 (64%) | 70/102 (69%) | 278/439 (63%) |
| Loss of appetite | | 249/541 (46%) | 66/102 (65%) | 183/439 (42%) |
| Abdominal pain or distension | | 89/541 (16%) | 24/102 (24%) | 65/439 (15%) |
| Peripheral lymph node swelling | | 6/541 (1%) | 4/102 (4%) | 2/439 (0%) |
| Test results | | | | |
| C-reactive protein (mg/l) | | 6 [3;24] (N=479) | 26 [10;58] (N=91) | 4 [3;17] (N=388) |
| C-reactive protein >5mg/l* | | 258/479 (54%) | 75/91 (82%) | 183/388 (47%) |
| CXR suggesting likely TB** | | 80/532 (15%) | 34/100 (34%) | 46/432 (11%) |
| CXR suggesting possible TB (includes likely)*** | | 263/532 (49%) | 81/100 (81%) | 182/432 (42%) |
| CXR suggestive of post-TB (with or without signs of active TB) | | 143/527 (27%) | 25/97 (26%) | 118/430 (27%) |
| TB-PCR on urine positive | | 6/519 (1%) | 6/101 (6%) | 0/418 (0%) |
| Number of sputa investigated for TB (<2; ≥2 samples) | | 45/541 (8%); 496/541 (92%) | 13/102 (13%); 89/102 (87%) | 32/439 (7%); 407/439 (93%) |
| Sputum smear status (% where done) | Negative | 488/507 (96) | 78/94 (83) | 410/413 (99) |
|  | scanty | 3/507 (1) | 3/94 (3) | 0/413 |
|  | 1+ | 4/507 (1) | 3/94 (3) | 1/413 (<1) |
|  | 2+ | 8/507 (2) | 6/94 (6) | 2/413 (<1) |
|  | 3+ | 4/507 (1) | 4/94 (4) | 0/413 |
|  | not done | 25 | 4 | 21 |
| Number of patients for whom bronchoalveolar lavage was investigated for TB | | 157/541 (29%) | 30/102 (29%) | 127/439 (29%) |
| Number of patients for whom non-sputum/non-BAL samples were investigated for TB | | 206/541 (38%) | 65/102 (64%) | 141/439 (32%) |
| Positive TB-PCR or culture on sputum or BAL | | 51/541 (9%) | 51/102 (50%) | 0/439 (0%) |
| Positive TB-PCR or culture on non-sputum/non-BAL sample | | 25/541 (5%) | 25/102 (25%) | 0/439 (0%) |
| PTB only  EPTB only  concurrent PTB+EPTB (%) | | NA | 51/102 (50)  17/102 (17)  34/102 (33) | NA |

**Legend**

denominators provided for all individuals with available data for each line.

* sensitivity 82% (95% confidence interval (CI) 73-89), specificity 53% (95% confidence interval 48-58)

** sensitivity 34% (95%-CI 25-44); specificity 89% (95%-CI 86-92)

*** sensitivity 81% (95%-CI 72-87); specificity 58% (95%-CI 53-62)

IQR, interquartile range;

n, number;

MRS, microbiological reference standard;

eMRS, extended MRS;

CRS, composite reference standard;

TB, tuberculoisis;

WHO, World Health Organization;

HIV, human immunodeficiency virus;

ART, anti-retroviral therapy;

CXR, chest x-ray;

PCR, polymerase chain reaction;

BAL, broncho-alveolar lavage;

EPTB, extra-pulmonary tuberculosis

NA, not applicable

| **Table 3 - POCUS – lung ultrasound, FASH, exploratory targets** | | | | | |
| --- | --- | --- | --- | --- | --- |
|  | **All participants (n=541)** | **CRS positive (n=102)** | **Unlikely TB (n=439)** | **Sensitivity (95%-CI)** | **Specificity (95%-CI)** |
| FASH | | | | | |
| FASH_original_ | 182/541 (34%) | 52/102 (51%) | 130/439 (30%) | 0·51 [0·41;0·6] | 0·7 [0·66;0·74] |
| FASH_ascites_ | 187/541 (35%) | 53/102 (52%) | 134/439 (31%) | 0·52 [0·42;0·61] | 0·69 [0·65;0·74] |
| FASH_pericardium_ | 196/541 (36%) | 54/102 (53%) | 142/439 (32%) | 0·53 [0·43;0·62] | 0·68 [0·63;0·72] |
| FASH_pleura600ml_ | 105/541 (19%) | 36/102 (35%) | 69/439 (16%) | 0·35 [0·27;0·45] | 0·84 [0·81;0·87] |
| Pleural effusion present, any | 172/541 (32%) | 47/102 (46%) | 125/439 (28%) | 0·46 [0·37;0·56] | 0·72 [0·67;0·76] |
| Pleural effusion volume estimate | 637 [288;1262] (N=172) | 914 [513;1483] (N=47) | 462 [256;1160] (N=125) | NA | NA |
| Pericardial effusion ≥10mm | 8/541 (1%) | 2/102 (2%) | 6/439 (1%) | 0·02 [0·01;0·07] | 0·99 [0·97;0·99] |
| Hypoechoic spleen lesions <1·5cm present | 7/541 (1%) | 4/102 (4%) | 3/439 (1%) | 0·04 [0·02;0·1] | 0·99 [0·98;1] |
| Hyperechoic spleen lesions present with or without calcification | 23/540 (4%) | 5/102 (5%) | 18/438 (4%) | NA | NA |
| Hypoechoic liver lesions | 5/541 (1%) | 0/102 (0%) | 5/439 (1%) | 0 [0;0·04] | 0·99 [0·97;1] |
| Abdominal lymph nodes ≥1·5cm present | 15/541 (3%) | 7/102 (7%) | 8/439 (2%) | 0·07 [0·03;0·13] | 0·98 [0·96;0·99] |
| Ascites present | 30/541 (6%) | 14/102 (14%) | 16/439 (4%) | 0·14 [0·08;0·22] | 0·96 [0·94;0·98] |
| LUNG ULTRASOUND | | | | | |
| Subpleural consolidations (SPC) <1cm present | 464/541 (86%) | 95/102 (93%) | 369/439 (84%) | 0·93 [0·87;0·97] | 0·16 [0·13;0·2] |
| SPCs with regular round/oval shape, hypoechoic echogenicity and posterior enhancement | 12/541 (2%) | 1/102 (1%) | 11/439 (3%) | 0·01 [0;0·05] | 0·97 [0·96;0·99] |
| >5 SPCs_<1cm_ with at least one ≥5mm | 150/541 (28%) | 32/102 (31%) | 118/439 (27%) | 0·31 [0·23;0·41] | 0·73 [0·69;0·77] |
| Subpleural consolidations ≥1cm present | 272/541 (50%) | 73/102 (72%) | 199/439 (45%) | 0·72 [0·62;0·79] | 0·55 [0·5;0·59] |
| any subpleural consolidation present, regardless of size | 476/541 (88%) | 100/102 (98%) | 376/439 (86%) | 0·98 [0·93;0·99] | 0·14 [0·11;0·18] |
| any consolidation in the apical regions | 294/541 (54%) | 66/102 (65%) | 228/439 (52%) | 0·65 [0·55;0·73] | 0·48 [0·43;0·53] |
| any consolidations <1cm in the apical regions | 252/541 (47%) | 57/102 (56%) | 195/439 (44%) | 0·56 [0·46;0·65] | 0·56 [0·51;0·6] |
| any consolidations ≥1cm in the apical regions | 84/541 (16%) | 22/102 (22%) | 62/439 (14%) | 0·22 [0·15;0·31] | 0·86 [0·82;0·89] |
| miliary pattern present | 63/541 (12%) | 13/102 (13%) | 50/439 (11%) | 0·13 [0·08;0·21] | 0·89 [0·85;0·91] |
| LUS findings in at least … lung zones | | | | | |
| SPC_<1cm_ in at least 1 lung zone | 464/541 (86%) | 95/102 (93%) | 369/439 (84%) | 0·93 [0·87;0·97] | 0·16 [0·13;0·2] |
| ≥2 lung zones | 390/541 (72%) | 84/102 (82%) | 306/439 (70%) | 0·82 [0·74;0·89] | 0·3 [0·26;0·35] |
| ≥3 lung zones | 315/541 (58%) | 72/102 (71%) | 243/439 (55%) | 0·71 [0·61;0·79] | 0·45 [0·4;0·49] |
| ≥4 lung zones | 247/541 (46%) | 52/102 (51%) | 195/439 (44%) | 0·51 [0·41;0·6] | 0·56 [0·51;0·6] |
| ≥5 lung zones | 186/541 (34%) | 38/102 (37%) | 148/439 (34%) | 0·37 [0·28;0·47] | 0·66 [0·62;0·71] |
| ≥6 lung zones | 146/541 (27%) | 22/102 (22%) | 124/439 (28%) | 0·22 [0·15;0·31] | 0·72 [0·67;0·76] |
| ≥7 lung zones | 114/541 (21%) | 16/102 (16%) | 98/439 (22%) | 0·16 [0·1;0·24] | 0·78 [0·74;0·81] |
| ≥8 lung zones | 83/541 (15%) | 15/102 (15%) | 68/439 (15%) | 0·15 [0·09;0·23] | 0·85 [0·81;0·88] |
| ≥9 lung zones | 53/541 (10%) | 11/102 (11%) | 42/439 (10%) | 0·11 [0·06;0·18] | 0·9 [0·87;0·93] |
| SPC_≥1cm_ in at least 1 lung zone | 272/541 (50%) | 73/102 (72%) | 199/439 (45%) | 0·72 [0·62;0·79] | 0·55 [0·5;0·59] |
| ≥2 lung zones | 194/541 (36%) | 57/102 (56%) | 137/439 (31%) | 0·56 [0·46;0·65] | 0·69 [0·64;0·73] |
| ≥3 lung zones | 129/541 (24%) | 39/102 (38%) | 90/439 (21%) | 0·38 [0·29;0·48] | 0·79 [0·75;0·83] |
| ≥4 lung zones | 86/541 (16%) | 27/102 (26%) | 59/439 (13%) | 0·26 [0·19;0·36] | 0·87 [0·83;0·89] |
| ≥5 lung zones | 52/541 (10%) | 13/102 (13%) | 39/439 (9%) | 0·13 [0·08;0·21] | 0·91 [0·88;0·93] |
| B-lines in at least 1 lung zone | 246/541 (45%) | 53/102 (52%) | 193/439 (44%) | 0·52 [0·42;0·61] | 0·56 [0·51;0·61] |
| ≥2 lung zones | 176/541 (33%) | 37/102 (36%) | 139/439 (32%) | 0·36 [0·28;0·46] | 0·68 [0·64;0·73] |
| ≥3 lung zones | 120/541 (22%) | 24/102 (24%) | 96/439 (22%) | 0·24 [0·16;0·33] | 0·78 [0·74;0·82] |
| ≥4 lung zones | 92/541 (17%) | 20/102 (20%) | 72/439 (16%) | 0·2 [0·13;0·28] | 0·84 [0·8;0·87] |
| ≥5 lung zones | 72/541 (13%) | 15/102 (15%) | 57/439 (13%) | 0·15 [0·09;0·23] | 0·87 [0·84;0·9] |
| ≥6 lung zones | 51/541 (9%) | 10/102 (10%) | 41/439 (9%) | 0·1 [0·05;0·17] | 0·91 [0·88;0·93] |
| Other targets | | | | | |
| IMNs ≥0·5cm present | 34/541 (6%) | 16/102 (16%) | 18/439 (4%) | 0·16 [0·1;0·24] | 0·96 [0·94;0·97] |
| Pleural thickening | 44/541 (8%) | 14/102 (14%) | 30/439 (7%) | NA | NA |
| Intestinal thickening in the right lower quadrant >4mm | 15/530 (3%) | 9/102 (9%) | 6/428 (1%) | 0·09 [0·05;0·16] | 0·99 [0·97;0·99] |
| Any peritoneal thickening * | 14/541 (3%) | 11/102 (11%) | 3/439 (1%) | 0·11 [0·06;0·18] | 0·99 [0·98;1] |
| Mediastinal lymph nodes seen from suprasternal view ** | 2/540 (0%) | 0/102 (0%) | 2/438 (0%) | NA | NA |
| Peripheral lymph nodes present (only if clinical suspicion) | 3/539 (1%) | 1/102 (1%) | 2/437 (0%) | NA | NA |
| **Legend**  denominators provided for all individuals with available data for each line.  * This affected the omentum in most cases (10/11) with additional hypoechoic nodules in 8/10  ** parasternal only negative  IQR, interquartile range;  n, number;  MRS, microbiological reference standard;  eMRS, extended MRS;  CRS, composite reference standard;  TB, tuberculoisis;  FASH, focused assessment with sonography for HIV-associated tuberculosis;  SPC, subpleural consolidation;  LUS, lung ultrasound;  IMN, internal mammars lymph node  NA, not applicable | | | | | |

| **Table 4 - POCUS – stratified by diabetes** | | | | | | |
| --- | --- | --- | --- | --- | --- | --- |
|  | DM-, all  (n=391) | DM-, CRS+ (n=71) | DM-, unlikely (n=320) | DM+, all (n=121) | DM+, CRS+ (n=23) | DM+, unlikely (n=98) |
| CXR suggestive of active TB | 54/383 (14%) | 24/69 (35%) | 30/314 (10%) | 24/121 (20%) | 9/23 (39%) | 15/98 (15%) |
| CXR suggestive or consistent with active TB | 180/383 (47%) | 56/69 (81%) | 124/314 (39%) | 74/121 (61%) | 20/23 (87%) | 54/98 (55%) |
| Positive TB-PCR on sputum or BAL | 32/391 (8%) | 32/71 (45%) | 0/320 (0%) | 14/121 (12%) | 14/23 (61%) | 0/98 (0%) |
| Positive TB-culture on sputum or BAL | 28/391 (7%) | 28/71 (39%) | 0/320 (0%) | 10/121 (8%) | 10/23 (43%) | 0/98 (0%) |
| Positive TB-PCR on non-sputum/BAL sample | 13/391 (3%) | 13/71 (18%) | 0/320 (0%) | 4/121 (3%) | 4/23 (17%) | 0/98 (0%) |
| Positive TB-culture on non-sputum/BAL sample | 8/391 (2%) | 8/71 (11%) | 0/320 (0%) | 3/121 (2%) | 3/23 (13%) | 0/98 (0%) |
| PTB only  EPTB only  PTB+EPTB | 34/71 (48)  14/71 (20)  23/71 (32) | | | 14/23 (61)  1/23 (4)  8/23 (35) | | |
| FASH (original) positive | 129/391 (33%) | 39/71 (55%) | 90/320 (28%) | 45/121 (37%) | 10/23 (43%) | 35/98 (36%) |
| Pleural effusion present, any | 121/391 (31%) | 34/71 (48%) | 87/320 (27%) | 44/121 (36%) | 10/23 (43%) | 34/98 (35%) |
| Pericardial effusion ≥10mm | 5/391 (1%) | 2/71 (3%) | 3/320 (1%) | 2/121 (2%) | 0/23 (0%) | 2/98 (2%) |
| Hypoechoic spleen lesions <1·5cm present | 5/391 (1%) | 3/71 (4%) | 2/320 (1%) | 1/121 (1%) | 1/23 (4%) | 0/98 (0%) |
| Hypoechoic liver lesions | 4/391 (1%) | 0/71 (0%) | 4/320 (1%) | 1/121 (1%) | 0/23 (0%) | 1/98 (1%) |
| Abdominal lymph nodes ≥1·5cm present | 9/391 (2%) | 4/71 (6%) | 5/320 (2%) | 5/121 (4%) | 3/23 (13%) | 2/98 (2%) |
| Ascites present | 18/391 (5%) | 6/71 (8%) | 12/320 (4%) | 9/121 (7%) | 5/23 (22%) | 4/98 (4%) |
| Subpleural consolidations (SPC) <1cm present | 333/391 (85%) | 67/71 (94%) | 266/320 (83%) | 108/121 (89%) | 22/23 (96%) | 86/98 (88%) |
| Subpleural consolidations ≥1cm present | 195/391 (50%) | 52/71 (73%) | 143/320 (45%) | 65/121 (54%) | 17/23 (74%) | 48/98 (49%) |
| any subpleural consolidation present, regardless of size? | 340/391 (87%) | 70/71 (99%) | 270/320 (84%) | 112/121 (93%) | 23/23 (100%) | 89/98 (91%) |
| miliary pattern present | 47/391 (12%) | 11/71 (15%) | 36/320 (11%) | 13/121 (11%) | 1/23 (4%) | 12/98 (12%) |
| B-lines (>2) in at least one lung zone | 182/391 (47%) | 40/71 (56%) | 142/320 (44%) | 49/121 (40%) | 8/23 (35%) | 41/98 (42%) |
| Internal mammary lymph nodes (IMNs) ≥0·5cm present | 28/391 (7%) | 13/71 (18%) | 15/320 (5%) | 5/121 (4%) | 2/23 (9%) | 3/98 (3%) |
| Pleural nodules or laminar thickening | 30/391 (8%) | 11/71 (15%) | 19/320 (6%) | 12/121 (10%) | 3/23 (13%) | 9/98 (9%) |
| Intestinal thickening in the right lower quadrant >4mm | 9/385 (2%) | 6/71 (8%) | 3/314 (1%) | 2/117 (2%) | 1/23 (4%) | 1/94 (1%) |
| Any peritoneal thickening | 6/391 (2%) | 4/71 (6%) | 2/320 (1%) | 6/121 (5%) | 5/23 (22%) | 1/98 (1%) |
| Peripheral lymph nodes present (only if clinical suspicion) | 2/390 (1%) | 0/71 (0%) | 2/319 (1%) | 1/120 (1%) | 1/23 (4%) | 0/97 (0%) |

**Legend**

denominators provided for all individuals with available data for each line.

DM, diabetes mellitus;

n, number;

HIV, human immunodeficiency virus;

CRS, composite reference standard;

CXR, chest x-ray;

TB, tuberculosis;

PCR, polymerase chain reaction;

BAL, broncho-alveolar lavage;

EPTB, extra-pulmonary tuberculosis;

PTB, pulmonary tuberculosis;

FASH, focused assessment with sonography for HIV-associated tuberculosis;

SPC, subpleural consolidations;

IMN, internal mammary lymph nodes

| **Table 5 - Differential diagnoses** | | | | | | | | | | | | | | | | | | | | | | | | | | |
| --- | --- | --- | --- | --- | --- | --- | --- | --- | --- | --- | --- | --- | --- | --- | --- | --- | --- | --- | --- | --- | --- | --- | --- | --- | --- | --- |
| Positive findings | In TB cases (CRS+) n=102 | All unlikely TB | Non-TB lung infection | Asthma | COPD or other obstructive lung disease | Post-TB sequelae | Lung neoplasia | bronchiectasis | ILD | Post-infectious sequelae | Inflammatory bowel disease | Chronic kidney disease | Non-infectious, non-inflammatory gastrointestinal disorder | Systemic autoimmune disorders | Heart disease | Hematological malignancy | Non-pulmonary primary malignancy | Pulmonary arterial hypertension incl· embolic | Gastrointestinal infections | Liver cirrhosis | Cholecystitis | Appendicitis | Tonsillitis | Pulmonary echinococcosis | gingivitis | Pleural empyema |
| N unlikely TB | NA | 439 | 102 | 92 | 71 | 51 | 48 | 44 | 27 | 25 | 17 | 14 | 14 | 12 | 11 | 9 | 8 | 7 | 4 | 2 | 1 | 1 | 1 | 1 | 1 | 1 |
| FASH_original_ | 52 | 130 | 28 | 15 | 18 | 8 | 31 | 7 | 9 | 4 | 1 | 9 | 1 | 8 | 9 | 5 | 5 | 5 | 1 | 2 | 0 | 0 | 0 | 0 | 0 | 1 |
| Pleural effusion, any | 47 | 125 | 28 | 13 | 17 | 8 | 31 | 7 | 8 | 3 | 1 | 9 | 1 | 8 | 9 | 4 | 5 | 5 | 1 | 2 | 0 | 0 | 0 | 0 | 0 | 1 |
| Pleural effusion >400ml | 42 | 68 | 14 | 2 | 6 | 4 | 25 | 1 | 2 | 0 | 0 | 6 | 0 | 2 | 7 | 3 | 3 | 3 | 0 | 2 | 0 | 0 | 0 | 0 | 0 | 1 |
| Pericardial effusion >1cm | 2 | 6 | 1 | 1 | 0 | 0 | 3 | 0 | 0 | 0 | 0 | 1 | 0 | 0 | 0 | 0 | 1 | 0 | 0 | 0 | 0 | 0 | 0 | 0 | 0 | 0 |
| Hypoechoic spleen lesions | 4 | 3 | 0 | 0 | 1 | 0 | 0 | 0 | 1 | 0 | 0 | 1 | 0 | 1 | 0 | 2 | 0 | 0 | 0 | 0 | 0 | 0 | 0 | 0 | 0 | 0 |
| Liver lesions | 0 | 5 | 1 | 1 | 0 | 0 | 0 | 0 | 2 | 0 | 0 | 1 | 0 | 1 | 0 | 0 | 2 | 2 | 0 | 0 | 0 | 0 | 0 | 0 | 0 | 0 |
| Abdominal lymph nodes | 7 | 8 | 1 | 0 | 1 | 0 | 1 | 0 | 1 | 1 | 0 | 2 | 0 | 2 | 0 | 3 | 0 | 0 | 0 | 1 | 0 | 0 | 0 | 0 | 0 | 0 |
| Ascites | 14 | 16 | 3 | 1 | 3 | 1 | 1 | 1 | 0 | 0 | 1 | 6 | 2 | 1 | 2 | 1 | 1 | 1 | 0 | 1 | 0 | 0 | 0 | 0 | 0 | 0 |
| Internal mammary lymph nodes | 16 | 18 | 12 | 3 | 1 | 4 | 2 | 2 | 0 | 1 | 0 | 1 | 0 | 0 | 0 | 1 | 0 | 0 | 0 | 0 | 0 | 0 | 0 | 0 | 0 | 1 |
| Peritoneal thickening | 11 | 3 | 0 | 0 | 0 | 0 | 1 | 0 | 0 | 0 | 0 | 1 | 0 | 0 | 0 | 0 | 0 | 0 | 0 | 1 | 0 | 0 | 0 | 0 | 0 | 0 |
| SPC_<1cm_ | 95 | 369 | 96 | 71 | 67 | 49 | 43 | 42 | 26 | 22 | 8 | 13 | 9 | 11 | 10 | 9 | 8 | 6 | 4 | 2 | 1 | 0 | 1 | 1 | 1 | 1 |
| SPC_≥1cm_ | 73 | 199 | 67 | 24 | 32 | 38 | 35 | 20 | 19 | 10 | 0 | 9 | 3 | 6 | 6 | 7 | 5 | 4 | 1 | 1 | 0 | 0 | 0 | 1 | 1 | 1 |
| Miliary pattern | 13 | 50 | 15 | 4 | 11 | 11 | 7 | 2 | 14 | 2 | 0 | 3 | 0 | 2 | 3 | 2 | 1 | 3 | 0 | 0 | 0 | 0 | 0 | 0 | 0 | 0 |
| CXR suggesting likely TB | 34 | 46 | 24 | 4 | 5 | 7 | 5 | 7 | 7 | 3 | 0 | 0 | 0 | 1 | 2 | 3 | 0 | 1 | 0 | 2 | 0 | 0 | 0 | 0 | 0 | 1 |
| Legend  TB, tuberculosis;  n, number;  ILD, interstitial lung disease;  COPD, chronic obstructive pulmonary disease;  FASH, focused assessment with sonography for HIV-associated tuberculosis;  SPC, subpleural consolidations;  CXR, chest x-ray | | | | | | | | | | | | | | | | | | | | | | | | | | |
