## Supplemental data for "Accuracy of lung and abdominal ultrasound for tuberculosis diagnosis: a prospective cohort study from India"

Main Heading 12p bold

Headings 10p bold Times New Roman

Text 10p

Study protocol: DOI

SAP: DOI

Supplement Table 1 comparison of PP vs. ITT

Supplement Table 2 extended (ITT)

Supplement Table 3 extended (ITT)

Supplement Table 4 - POCUS – stratified by HIV and diabetes

Supplement Table 5 PP data

Interrater analysis

Regression analyses

STARD checklist

| **Supplement Table 1 Comparison of per protocol (PP) and intention to test (ITT) groups** | | |
| --- | --- | --- |
|  | **ITT** | **PP** |
| N | 541 | 489 |
| Age in years | 48 [35;59] (N=541) | 48 [35;60] (N=489) |
| Sex male | 357/541 (66%) | 324/489 (66%) |
| Body mass index (kg/m^2) | 21·6 [18·4;24·9] (N=541) | 21·7 [18·6;25·1] (N=489) |
| History of tobacco use | 110/541 (20%) | 95/489 (19%) |
| Pack years | 10 [3;22] (N=109) | 10 [3;22] (N=95) |
| History of diabetes | 110/541 (20%) | 104/489 (21%) |
| Final diabetes status | 121/512 (24%) | 113/462 (24%) |
| Diabetics using insulin | 19/121 (16%) | 17/113 (15%) |
| History of known HIV-infection | 5/357 (1%) | 5/327 (2%) |
| Patients already on ART | 2/5 (40%) | 2/5 (40%) |
| Final HIV status | 6/523 (1%) | 6/472 (1%) |
| History of previous COVID-19 | 50/541 (9%) | 47/489 (10%) |
| History of previous COVID-19 with hospitalization | 32/50 (64%) | 29/47 (62%) |
| History of previous TB disease | 99/540 (18%) | 87/488 (18%) |
| History of current TB-contact | 5/535 (1%) | 5/483 (1%) |
| History of previous TB-contact | 43/535 (8%) | 35/483 (7%) |
| CURRENT SYMPTOMS |  |  |
| Cough | 483/541 (89%) | 440/489 (90%) |
| duration of patients with cough | 12 [4;24] (N=483) | 12 [4;24] (N=440) |
| Hemoptysis | 134/541 (25%) | 109/489 (22%) |
| duration of patients with hemoptysis | 12 [4;24] (N=133) | 8 [4;16] (N=108) |
| Night sweats | 48/541 (9%) | 46/489 (9%) |
| duration of patients with night sweats | 8 [4;18] (N=47) | 8 [4;16] (N=45) |
| Fever | 230/541 (43%) | 207/489 (42%) |
| duration of patients with fever | 4 [4;12] (N=230) | 4 [4;12] (N=207) |
| Weight loss | 292/541 (54%) | 259/489 (53%) |
| duration of patients with weight loss | 12 [8;26] (N=292) | 12 [8;24] (N=259) |
| Fatigue | 348/541 (64%) | 316/489 (65%) |
| duration of patients with fatigue | 12 [4;24] (N=348) | 10 [4;24] (N=316) |
| Loss of appetite | 249/541 (46%) | 220/489 (45%) |
| duration of patients with loss of appetite | 10 [4;20] (N=249) | 8 [4;20] (N=220) |
| Abdominal pain or distension | 89/541 (16%) | 82/489 (17%) |
| duration of patients with abdminal pain or distension | 18 [8;54] (N=88) | 20 [8;54] (N=81) |
| Peripheral lymph node swelling | 6/541 (1%) | 6/489 (1%) |
| duration of patients with peripheral lymph node swelling | 8 [5;11] (N=6) | 8 [5;11] (N=6) |
| DIAGNOSTICS |  |  |
| HbA1c (%) | 6 [5;6] (N=510) | 6 [5;6] (N=460) |
| HBA1c (%) in diabetic patients | 7 [6;8] (N=119) | 7 [6;8] (N=111) |
| C-reactive protein (mg/l) | 6 [3;24] (N=479) | 6 [3;23] (N=437) |
| Hemoglobin (g/dl) | 12 [11;14] (N=515) | 12 [11;14] (N=463) |
| Platelet count (/nl) | 245 [176;330] (N=487) | 244 [181;328] (N=436) |
| White blood count (/nl) | 8 [6;10] (N=510) | 8 [6;10] (N=461) |
| CXR suggesting likely TB | 80/532 (15%) | 67/480 (14%) |
| CXR suggesting possible TB (including likely) | 263/532 (49%) | 231/480 (48%) |
| CXR suggestive of post-TB (with or without signs of active TB) | 143/527 (27%) | 129/476 (27%) |
| MRS+ | 59/541 (11%) | 54/489 (11%) |
| eMRS+ | 76/541 (14%) | 67/489 (14%) |
| CRS+ | 102/541 (19%) | 92/489 (19%) |
| Unlikely TB | 439/541 (81%) | 397/489 (81%) |
| **Legend**  IQR, interquartile range;  n, number;  MRS, microbiological reference standard;  eMRS, extended MRS;  CRS, composite reference standard;  TB, tuberculoisis;  WHO, World Health Organization;  HIV, human immunodeficiency virus;  ART, anti-retroviral therapy;  CXR, chest x-ray;  NA, not applicable | | |

| **Supplement Table 2 - patient characteristics and reference standard testing - extended** | | | | | | |
| --- | --- | --- | --- | --- | --- | --- |
| Variables in n, median, IQR, (%) | | All participants (n=541) | MRS positive (n=59) | eMRS positive (n=76) | CRS positive (n=102) | Unlikely TB (n=439) |
| Age in years | | 48 [35;59] (N=541) | 47 [29;57] (N=59) | 46 [30;57] (N=76) | 44 [29;56] (N=102) | 49 [36;60] (N=439) |
| Sex male | | 357/541 (66%) | 37/59 (63%) | 47/76 (62%) | 65/102 (64%) | 292/439 (67%) |
| Body mass index (kg/m^2) | | 21·6 [18·4;24·9] (N=541) | 19 [16·8;22·3] (N=59) | 19 [16·6;22·5] (N=76) | 19·4 [17·1;23·1] (N=102) | 22 [19·1;25·2] (N=439) |
| History of tobacco use * | | 110/541 (20%) | 11/59 (19%) | 12/76 (16%) | 18/102 (18%) | 92/439 (21%) |
| Pack years | | 10 [3;22] (N=109) | 4 [2;26] (N=11) | 7 [2;32] (N=12) | 10 [3;22] (N=18) | 10 [3;22] (N=91) |
| History of diabetes | | 110/541 (20%) | 16/59 (27%) | 19/76 (25%) | 22/102 (22%) | 88/439 (20%) |
| Final diabetes status | | 121/512 (24%) | 16/56 (29%) | 19/71 (27%) | 23/94 (24%) | 98/418 (23%) |
| Diabetics using insulin | | 19/121 (16%) | 1/16 (6%) | 2/19 (11%) | 2/23 (9%) | 17/98 (17%) |
| History of known HIV-infection | | 5/357 (1%) | 1/41 (2%) | 1/51 (2%) | 1/66 (2%) | 4/291 (1%) |
| Patients already on ART | | 2/5 (40%) | 0/1 (0%) | 0/1 (0%) | 0/1 (0%) | 2/4 (50%) |
| Final HIV status | | 6/523 (1%) | 1/58 (2%) | 1/75 (1%) | 1/101 (1%) | 5/422 (1%) |
| History of previous COVID-19 | | 50/541 (9%) | 4/59 (7%) | 4/76 (5%) | 8/102 (8%) | 42/439 (10%) |
| History of previous COVID-19 with hospitalization | | 32/50 (64%) | 2/4 (50%) | 2/4 (50%) | 6/8 (75%) | 26/42 (62%) |
| History of previous TB disease | | 99/540 (18%) | 7/59 (12%) | 10/76 (13%) | 11/102 (11%) | 88/438 (20%) |
| History of current TB-contact | | 5/535 (1%) | 1/59 (2%) | 1/76 (1%) | 3/102 (3%) | 2/433 (0%) |
| History of previous TB-contact | | 43/535 (8%) | 10/59 (17%) | 10/76 (13%) | 13/102 (13%) | 30/433 (7%) |
| Symptoms | | | | | | |
| Cough | | 483/541 (89%) | 55/59 (93%) | 68/76 (89%) | 89/102 (87%) | 394/439 (90%) |
| duration of patients with cough | | 12 [4;24] (N=483) | 12 [4;24] (N=55) | 12 [6;24] (N=68) | 8 [4;24] (N=89) | 12 [4;24] (N=394) |
| Hemoptysis | | 134/541 (25%) | 11/59 (19%) | 15/76 (20%) | 18/102 (18%) | 116/439 (26%) |
| duration of patients with hemoptysis | | 12 [4;24] (N=133) | 16 [3;26] (N=11) | 12 [3;25] (N=15) | 12 [2;23] (N=18) | 12 [4;24] (N=115) |
| Night sweats | | 48/541 (9%) | 10/59 (17%) | 12/76 (16%) | 16/102 (16%) | 32/439 (7%) |
| duration of patients with night sweats | | 8 [4;18] (N=47) | 4 [2;4] (N=10) | 4 [4;5] (N=12) | 4 [4;8] (N=16) | 12 [4;25] (N=31) |
| Fever | | 230/541 (43%) | 42/59 (71%) | 53/76 (70%) | 68/102 (67%) | 162/439 (37%) |
| duration of patients with fever | | 4 [4;12] (N=230) | 4 [4;12] (N=42) | 4 [4;10] (N=53) | 4 [4;12] (N=68) | 4 [4;12] (N=162) |
| Weight loss | | 292/541 (54%) | 42/59 (71%) | 54/76 (71%) | 74/102 (73%) | 218/439 (50%) |
| duration of patients with weight loss | | 12 [8;26] (N=292) | 8 [4;26] (N=42) | 9 [4;24] (N=54) | 11 [4;24] (N=74) | 12 [8;26] (N=218) |
| Fatigue | | 348/541 (64%) | 40/59 (68%) | 53/76 (70%) | 70/102 (69%) | 278/439 (63%) |
| duration of patients with fatigue | | 12 [4;24] (N=348) | 9 [4;21] (N=40) | 8 [4;20] (N=53) | 8 [4;20] (N=70) | 12 [4;24] (N=278) |
| Loss of appetite | | 249/541 (46%) | 36/59 (61%) | 47/76 (62%) | 66/102 (65%) | 183/439 (42%) |
| duration of patients with loss of appetite | | 10 [4;20] (N=249) | 8 [4;13] (N=36) | 8 [4;16] (N=47) | 8 [4;19] (N=66) | 12 [4;24] (N=183) |
| Abdominal pain or distension | | 89/541 (16%) | 9/59 (15%) | 13/76 (17%) | 24/102 (24%) | 65/439 (15%) |
| duration of patients with abdominal pain or distension | | 18 [8;54] (N=88) | 16 [12;52] (N=9) | 12 [4;48] (N=13) | 16 [8;37] (N=24) | 22 [8;81] (N=64) |
| Peripheral lymph node swelling | | 6/541 (1%) | 1/59 (2%) | 3/76 (4%) | 4/102 (4%) | 2/439 (0%) |
| duration of patients with peripheral lymph node swelling | | 8 [5;11] (N=6) | 8 [8;8] (N=1) | 8 [6;8] (N=3) | 8 [7;9] (N=4) | 13 [8;18] (N=2) |
| Test results | | | | | | |
| HbA1c (%) | | 6 [5;6] (N=510) | 6 [5;6] (N=55) | 6 [5;6] (N=70) | 6 [5;6] (N=93) | 6 [5;6] (N=417) |
| HBA1c (%) in diabetic patients | | 7 [6;8] (N=119) | 7 [6;9] (N=15) | 8 [7;9] (N=18) | 7 [6;9] (N=22) | 7 [6;8] (N=97) |
| C-reactive protein (mg/l) | | 6 [3;24] (N=479) | 26 [6;60] (N=51) | 27 [7;58] (N=66) | 26 [10;58] (N=91) | 4 [3;17] (N=388) |
| C-reactive protein >5mg/l* | | 258/479 (54%) | 39/51 (76%) | 53/66 (80%) | 75/91 (82%) | 183/388 (47%) |
| Hemoglobin (g/dl) | | 12 [11;14] (N=515) | 11 [11;13] (N=55) | 11 [10;13] (N=72) | 11 [10;13] (N=98) | 12 [11;14] (N=417) |
| Platelet count (/nl) | | 245 [176;330] (N=487) | 311 [214;381] (N=53) | 282 [189;378] (N=70) | 294 [200;366] (N=95) | 234 [174;314] (N=392) |
| White blood count (/nl) | | 8 [6;10] (N=510) | 7 [6;8] (N=52) | 7 [6;8] (N=68) | 7 [6;9] (N=93) | 8 [6;10] (N=417) |
| CXR suggesting likely TB** | | 80/532 (15%) | 23/57 (40%) | 29/74 (39%) | 34/100 (34%) | 46/432 (11%) |
| CXR suggesting possible TB (includes likely)*** | | 263/532 (49%) | 48/57 (84%) | 62/74 (84%) | 81/100 (81%) | 182/432 (42%) |
| CXR suggestive of post-TB (with or without signs of active TB) | | 143/527 (27%) | 19/56 (34%) | 22/72 (31%) | 25/97 (26%) | 118/430 (27%) |
| TB-PCR on urine positive | | 6/519 (1%) | 6/59 (10%) | 6/75 (8%) | 6/101 (6%) | 0/418 (0%) |
| Number of sputa investigated for TB (<2; ≥2 samples) | | 45/541 (8%); 496/541 (92%) | 8/59 (14%); 51/59 (86%) | 12/76 (16%); 64/76 (84%) | 13/102 (13%); 89/102 (87%) | 32/439 (7%); 407/439 (93%) |
| Sputum smear status (% where done) | Negative | 488/507 (96) | 38/54 (70) | 54/70 (77) | 78/94 (83) | 410/413 (99) |
|  | scanty | 3/507 (1) | 3/54 (6) | 3/70 (4) | 3/94 (3) | 0/413 |
|  | 1+ | 4/507 (1) | 3/54 (6) | 3/70 (4) | 3/94 (3) | 1/413 (<1) |
|  | 2+ | 8/507 (2) | 6/54 (11) | 6/70 (9) | 6/94 (6) | 2/413 (<1) |
|  | 3+ | 4/507 (1) | 4/54 (7) | 4/70 (6) | 4/94 (4) | 0/413 |
|  | not done | 25 | 3 | 3 | 4 | 21 |
| Number of patients for whom bronchoalveolar lavage was investigated for TB | | 157/541 (29%) | 23/59 (39%) | 26/76 (34%) | 30/102 (29%) | 127/439 (29%) |
| Number of patients for whom non-sputum/non-BAL samples were investigated for TB | | 206/541 (38%) | 24/59 (41%) | 41/76 (54%) | 65/102 (64%) | 141/439 (32%) |
| Positive TB-PCR or culture on sputum or BAL | | 51/541 (9%) | 51/59 (86%) | 51/76 (67%) | 51/102 (50%) | 0/439 (0%) |
| Positive TB-PCR or culture on non-sputum/BAL sample | | 25/541 (5%) | 8/59 (14%) | 25/76 (33%) | 25/102 (25%) | 0/439 (0%) |
| PTB only  EPTB only  concurrent PTB+EPTB (%) | | NA | 45/59 (76)  2/59 (3)  12/59 (20) | 49/76 (64)  5/76 (7)  22/76 (29) | 51/102 (50)  17/102 (17)  34/102 (33) | NA |
| **Legend**  denominators provided for all individuals with available data for each line·  * sensitivity 82% (95% confidence interval (CI) 73-89), specificity 53% (95% confidence interval 48-58)  ** sensitivity 34% (95%-CI 25-44); specificity 89% (95%-CI 86-92)  *** sensitivity 81% (95%-CI 72-87); specificity 58% (95%-CI 53-62)  IQR, interquartile range;  n, number;  MRS, microbiological reference standard;  eMRS, extended MRS;  CRS, composite reference standard;  TB, tuberculoisis;  WHO, World Health Organization;  HIV, human immunodeficiency virus;  ART, anti-retroviral therapy;  CXR, chest x-ray;  PCR, polymerase chain reaction;  BAL, broncho-alveolar lavage;  EPTB, extra-pulmonary tuberculosis  NA, not applicable | | | | | | |

| **Table 3 – POCUS – lung ultrasound, FASH, exploratory targets – extended** | | | | | |
| --- | --- | --- | --- | --- | --- |
|  | **All participants (n=541)** | **CRS positive (n=102)** | **Unlikely TB (n=439)** | **Sensitivity (95%-CI)** | **Specificity (95%-CI)** |
| FASH | | | | | |
| FASH_original_ | 182/541 (34%) | 52/102 (51%) | 130/439 (30%) | 0·51 [0·41;0·6] | 0·7 [0·66;0·74] |
| FASH_ascites_ | 187/541 (35%) | 53/102 (52%) | 134/439 (31%) | 0·52 [0·42;0·61] | 0·69 [0·65;0·74] |
| FASH_pericardium_ | 196/541 (36%) | 54/102 (53%) | 142/439 (32%) | 0·53 [0·43;0·62] | 0·68 [0·63;0·72] |
| FASH_pleura200ml_ | 161/541 (30%) | 49/102 (48%) | 112/439 (26%) | 0·48 [0·39;0·58] | 0·74 [0·7;0·78] |
| FASH_pleura300ml_ | 137/541 (25%) | 44/102 (43%) | 93/439 (21%) | 0·43 [0·34;0·53] | 0·79 [0·75;0·82] |
| FASH_pleura400ml_ | 120/541 (22%) | 42/102 (41%) | 78/439 (18%) | 0·41 [0·32;0·51] | 0·82 [0·78;0·86] |
| FASH_pleura600ml_ | 105/541 (19%) | 36/102 (35%) | 69/439 (16%) | 0·35 [0·27;0·45] | 0·84 [0·81;0·87] |
| FASH_pleura800ml_ | 94/541 (17%) | 33/102 (32%) | 61/439 (14%) | 0·32 [0·24;0·42] | 0·86 [0·83;0·89] |
| FASH_pleura1000ml_ | 81/541 (15%) | 29/102 (28%) | 52/439 (12%) | 0·28 [0·21;0·38] | 0·88 [0·85;0·91] |
| Pleural effusion present, any | 172/541 (32%) | 47/102 (46%) | 125/439 (28%) | 0·46 [0·37;0·56] | 0·72 [0·67;0·76] |
| Pleural effusion echogenicity | Anechoic: 92/172 (53)  Mixed: 80/172 (47) | 25/47 (53)  22/47 (47) | 67/125 (54)  58/125 (46) | NA | NA |
| Pleural effusion volume estimate | 637 [288;1262] (N=172) | 914 [513;1483] (N=47) | 462 [256;1160] (N=125) | NA | NA |
| Pericardial effusion ≥4mm | 40/541 (7%) | 9/102 (9%) | 31/439 (7%) | 0·09 [0·05;0·16] | 0·93 [0·9;0·95] |
| Pericardial effusion ≥10mm | 8/541 (1%) | 2/102 (2%) | 6/439 (1%) | 0·02 [0·01;0·07] | 0·99 [0·97;0·99] |
| Pericardial effusion echogenicity | Anechoic: 29/40 (73)  Mixed: 11/40 (28) | Anechoic: 8/9 (89)  Mixed: 1/9 (11) | Anechoic: 21/31 (68)  Mixed: 10/31 (32) | NA | NA |
| Hypoechoic spleen lesions <1·5cm present | 7/541 (1%) | 4/102 (4%) | 3/439 (1%) | 0·04 [0·02;0·1] | 0·99 [0·98;1] |
| Number of hypoechoic spleen lesions | 2-5: 2/7 (29)  >5: 5 (71) | 2-5: ¼ (25)  >5: ¾ (75) | 2-5: 1/3 (33)  >5: 2/3 (67) | NA | NA |
| Larger splenic lesions present (≥1·5cm) suggestive of macroabscess | 3/540 (1%) | 2/102 (2%) | 1/438 (0%) | NA | NA |
| Hyperechoic spleen lesions present with or without calcification | 23/540 (4%) | 5/102 (5%) | 18/438 (4%) | NA | NA |
| Size of spleen | 10 [9;11] (N=530) | 10 [9;11] (N=102) | 10 [9;11] (N=428) | NA | NA |
| Hypoechoic liver lesions | 5/541 (1%) | 0/102 (0%) | 5/439 (1%) | 0 [0;0·04] | 0·99 [0·97;1] |
| Number of hypoechoic liver lesions | single: 2/5 (40)  2-5: 1/5 (20)  >5: 2/5 (40) | - | single: 2/5 (40)  2-5: 1/5 (20)  >5: 2/5 (40) | NA | NA |
| Abdominal lymph nodes ≥1·5cm present | 15/541 (3%) | 7/102 (7%) | 8/439 (2%) | 0·07 [0·03;0·13] | 0·98 [0·96;0·99] |
| Maximum size of abdominal lymph nodes | 2 [2;2] (N=15) | 2 [2;2] (N=7) | 2 [2;3] (N=8) | NA | NA |
| Number of abdominal lymph nodes | single: 7/15 (47)  2-5: 5/15 (33)  >5: 3/15 (20) | single: 3/7 (43)  2-5: 3/7 (43)  >5: 1/7 (14) | single: 4/8 (50)  2-5: 2/8 (25)  >5: 2/8 (25) | NA | NA |
| Additional morphological pathologies seen | Hilum loss: 1/15 (7)  bulking: 3/15 (20) | Hilum loss: 1/7 (14)  bulking: 1/7 (14) | Bulking: 2/8 (25) | NA | NA |
| Ascites present | 30/541 (6%) | 14/102 (14%) | 16/439 (4%) | 0·14 [0·08;0·22] | 0·96 [0·94;0·98] |
| Ascites amount | small: 10/30 (33)  moderate: 17/30 (57)  large: 3/30 (10) | small: 5/14 (36)  moderate: 8/14 (57)  large: 1/14 (7) | small: 5/16 (31)  moderate: 9/16 (56)  large: 2/16 (13) | NA | NA |
| Ascites echogenic quality | anechoic: 24/30 (80)  mixed: 6/30 (20) | anechoic: 11/14 (79)  mixed: 3/14 (21) | anechoic: 13/16 (81)  mixed: 3/16 (19) | NA | NA |
| LUNG ULTRASOUND | | | | | |
| <1cm consolidations | | | | | |
| Subpleural consolidations (SPC) <1cm present | 464/541 (86%) | 95/102 (93%) | 369/439 (84%) | 0·93 [0·87;0·97] | 0·16 [0·13;0·2] |
| SPCs with regular round/oval shape, hypoechoic echogenicity and posterior enhancement | 12/541 (2%) | 1/102 (1%) | 11/439 (3%) | 0·01 [0;0·05] | 0·97 [0·96;0·99] |
| Number of subpleural consolidations <1cm | single: 67/464 (14)  2-5: 227/464 (49)  >5: 170/464 (37) | single: 9/95 (9); 2-5: 49/95 (52); >5: 37/95 (39) | single: 58/369 (16); 2-5: 178/369 (48); >5: 133/369 (36) | NA | NA |
| Number of lung zones with subpleural consolidations <1cm | 4 [2;6] (N=464) | 4 [3;5] (N=95) | 4 [2;7] (N=369) | NA | NA |
| >5 SPCs present | 170/541 (31%) | 37/102 (36%) | 133/439 (30%) | 0·36 [0·28;0·46] | 0·7 [0·65;0·74] |
| SPCs with ≥5mm present | 355/541 (66%) | 75/102 (74%) | 280/439 (64%) | 0·74 [0·64;0·81] | 0·36 [0·32;0·41] |
| >5 SPCs with at least one ≥5mm | 150/541 (28%) | 32/102 (31%) | 118/439 (27%) | 0·31 [0·23;0·41] | 0·73 [0·69;0·77] |
| ≥1cm consolidations | | | | | |
| Subpleural consolidations ≥1cm present | 272/541 (50%) | 73/102 (72%) | 199/439 (45%) | 0·72 [0·62;0·79] | 0·55 [0·5;0·59] |
| Type of subpleural consolidations ≥1cm | translobar: 37/272 (14)  shred: 158/272 (58)  both: 77/272 (28) | translobar: 9/73 (12)  shred: 42/73 (58)  both: 22/73 (30) | translobar: 28/199 (14)  shred: 116/199 (58)  both: 55/199 (28) | NA | NA |
| bronchograms visualized? | Air: 135/272 (50)  fluid: 1/272 (<1)  air+fluid: 11/272 (4) | air: 44/73 (60)  air+fluid: 2/73 (3) | air: 91/199 (48)  fluid: 1/199 (1)  air+fluid: 9/199 (5) | NA | NA |
| Linear aerobronchograms visualized? | 61/147 (41%) | 21/46 (46%) | 40/101 (40%) | 0·41 [0·28;0·56] | 0·64 [0·55;0·73] |
| Number of lung zones with subpleural consolidations ≥1cm | 2 [1;4] (N=272) | 3 [2;4] (N=73) | 2 [1;4] (N=199) | NA | NA |
| combinations of <1cm and ≥1cm | | | | | |
| any subpleural consolidation present, regardless of size? | 476/541 (88%) | 100/102 (98%) | 376/439 (86%) | 0·98 [0·93;0·99] | 0·14 [0·11;0·18] |
| maximum size of consolidations, regardless of <1cm or ≥1cm | 1 [0;2] (N=472) | 1 [1;2] (N=100) | 1 [0;2] (N=372) | NA | NA |
| any subpleural consolidation present, size ≥5mm | 412/541 (76%) | 93/102 (91%) | 319/439 (73%) | 0·91 [0·84;0·95] | 0·27 [0·23;0·32] |
| Size of largest subpleural consolidation, categorized by small, medium, large | large: 118/476 (25)  medium: 154/476 (32)  small: 204/476 (43) | large: 35/100 (35)  medium: 38/100 (38)  small: 27/100 (27) | large: 83/376 (22)  medium: 116/376 (31)  small: 177/376 (47) | NA | NA |
| Location of consolidations | | | | | |
| any consolidation in the apical regions | 294/541 (54%) | 66/102 (65%) | 228/439 (52%) | 0·65 [0·55;0·73] | 0·48 [0·43;0·53] |
| any consolidations in the upper lung regions | 435/541 (80%) | 97/102 (95%) | 338/439 (77%) | 0·95 [0·89;0·98] | 0·23 [0·19;0·27] |
| any consolidations <1cm in the apical regions | 252/541 (47%) | 57/102 (56%) | 195/439 (44%) | 0·56 [0·46;0·65] | 0·56 [0·51;0·6] |
| any consolidations <1cm in the upper lung regions | 415/541 (77%) | 90/102 (88%) | 325/439 (74%) | 0·88 [0·81;0·93] | 0·26 [0·22;0·3] |
| any consolidations ≥1cm in the apical regions | 84/541 (16%) | 22/102 (22%) | 62/439 (14%) | 0·22 [0·15;0·31] | 0·86 [0·82;0·89] |
| any consollidations ≥1cm in the upper lung zones | 188/541 (35%) | 54/102 (53%) | 134/439 (31%) | 0·53 [0·43;0·62] | 0·69 [0·65;0·74] |
| Other LUS findings | | | | | |
| miliary pattern present | 63/541 (12%) | 13/102 (13%) | 50/439 (11%) | 0·13 [0·08;0·21] | 0·89 [0·85;0·91] |
| B-lines (>2) in at least one lung zone | 246/541 (45%) | 53/102 (52%) | 193/439 (44%) | 0·52 [0·42;0·61] | 0·56 [0·51;0·61] |
| Number of lung zones with B-lines | 2 [1;5] (N=246) | 2 [1;5] (N=53) | 2 [1;5] (N=193) | NA | NA |
| LUS findings in at least … lung zones | | | | | |
| SPC_<1cm_ in at least 1 lung zone | 464/541 (86%) | 95/102 (93%) | 369/439 (84%) | 0·93 [0·87;0·97] | 0·16 [0·13;0·2] |
| ≥2 lung zones | 390/541 (72%) | 84/102 (82%) | 306/439 (70%) | 0·82 [0·74;0·89] | 0·3 [0·26;0·35] |
| ≥3 lung zones | 315/541 (58%) | 72/102 (71%) | 243/439 (55%) | 0·71 [0·61;0·79] | 0·45 [0·4;0·49] |
| ≥4 lung zones | 247/541 (46%) | 52/102 (51%) | 195/439 (44%) | 0·51 [0·41;0·6] | 0·56 [0·51;0·6] |
| ≥5 lung zones | 186/541 (34%) | 38/102 (37%) | 148/439 (34%) | 0·37 [0·28;0·47] | 0·66 [0·62;0·71] |
| ≥6 lung zones | 146/541 (27%) | 22/102 (22%) | 124/439 (28%) | 0·22 [0·15;0·31] | 0·72 [0·67;0·76] |
| ≥7 lung zones | 114/541 (21%) | 16/102 (16%) | 98/439 (22%) | 0·16 [0·1;0·24] | 0·78 [0·74;0·81] |
| ≥8 lung zones | 83/541 (15%) | 15/102 (15%) | 68/439 (15%) | 0·15 [0·09;0·23] | 0·85 [0·81;0·88] |
| ≥9 lung zones | 53/541 (10%) | 11/102 (11%) | 42/439 (10%) | 0·11 [0·06;0·18] | 0·9 [0·87;0·93] |
| ≥10 lung zones | 38/541 (7%) | 9/102 (9%) | 29/439 (7%) | 0·09 [0·05;0·16] | 0·93 [0·91;0·95] |
| ≥11 lung zones | 27/541 (5%) | 6/102 (6%) | 21/439 (5%) | 0·06 [0·03;0·12] | 0·95 [0·93;0·97] |
| ≥12 lung zones | 17/541 (3%) | 4/102 (4%) | 13/439 (3%) | 0·04 [0·02;0·1] | 0·97 [0·95;0·98] |
| ≥13 lung zones | 9/541 (2%) | 2/102 (2%) | 7/439 (2%) | 0·02 [0·01;0·07] | 0·98 [0·97;0·99] |
| ≥14 lung zones | 6/541 (1%) | 1/102 (1%) | 5/439 (1%) | 0·01 [0;0·05] | 0·99 [0·97;1] |
| SPC_≥1cm_ in at least 1 lung zone | 272/541 (50%) | 73/102 (72%) | 199/439 (45%) | 0·72 [0·62;0·79] | 0·55 [0·5;0·59] |
| ≥2 lung zones | 194/541 (36%) | 57/102 (56%) | 137/439 (31%) | 0·56 [0·46;0·65] | 0·69 [0·64;0·73] |
| ≥3 lung zones | 129/541 (24%) | 39/102 (38%) | 90/439 (21%) | 0·38 [0·29;0·48] | 0·79 [0·75;0·83] |
| ≥4 lung zones | 86/541 (16%) | 27/102 (26%) | 59/439 (13%) | 0·26 [0·19;0·36] | 0·87 [0·83;0·89] |
| ≥5 lung zones | 52/541 (10%) | 13/102 (13%) | 39/439 (9%) | 0·13 [0·08;0·21] | 0·91 [0·88;0·93] |
| ≥6 lung zones | 32/541 (6%) | 6/102 (6%) | 26/439 (6%) | 0·06 [0·03;0·12] | 0·94 [0·91;0·96] |
| ≥7 lung zones | 22/541 (4%) | 5/102 (5%) | 17/439 (4%) | 0·05 [0·02;0·11] | 0·96 [0·94;0·98] |
| ≥8 lung zones | 11/541 (2%) | 4/102 (4%) | 7/439 (2%) | 0·04 [0·02;0·1] | 0·98 [0·97;0·99] |
| ≥9 lung zones | 5/541 (1%) | 1/102 (1%) | 4/439 (1%) | 0·01 [0;0·05] | 0·99 [0·98;1] |
| ≥10 lung zones | 2/541 (0%) | 0/102 (0%) | 2/439 (0%) | 0 [0;0·04] | 1 [0·98;1] |
| ≥11 lung zones | 1/541 (0%) | 0/102 (0%) | 1/439 (0%) | 0 [0;0·04] | 1 [0·99;1] |
| ≥12 lung zones | 0/541 (0%) | 0/102 (0%) | 0/439 (0%) | 0 [0;0·04] | 1 [0·99;1] |
| ≥13 lung zones | 0/541 (0%) | 0/102 (0%) | 0/439 (0%) | 0 [0;0·04] | 1 [0·99;1] |
| ≥14 lung zones | 0/541 (0%) | 0/102 (0%) | 0/439 (0%) | 0 [0;0·04] | 1 [0·99;1] |
| B-lines in at least 1 lung zone | 246/541 (45%) | 53/102 (52%) | 193/439 (44%) | 0·52 [0·42;0·61] | 0·56 [0·51;0·61] |
| ≥2 lung zones | 176/541 (33%) | 37/102 (36%) | 139/439 (32%) | 0·36 [0·28;0·46] | 0·68 [0·64;0·73] |
| ≥3 lung zones | 120/541 (22%) | 24/102 (24%) | 96/439 (22%) | 0·24 [0·16;0·33] | 0·78 [0·74;0·82] |
| ≥4 lung zones | 92/541 (17%) | 20/102 (20%) | 72/439 (16%) | 0·2 [0·13;0·28] | 0·84 [0·8;0·87] |
| ≥5 lung zones | 72/541 (13%) | 15/102 (15%) | 57/439 (13%) | 0·15 [0·09;0·23] | 0·87 [0·84;0·9] |
| ≥6 lung zones | 51/541 (9%) | 10/102 (10%) | 41/439 (9%) | 0·1 [0·05;0·17] | 0·91 [0·88;0·93] |
| ≥7 lung zones | 41/541 (8%) | 8/102 (8%) | 33/439 (8%) | 0·08 [0·04;0·15] | 0·92 [0·9;0·95] |
| ≥8 lung zones | 31/541 (6%) | 8/102 (8%) | 23/439 (5%) | 0·08 [0·04;0·15] | 0·95 [0·92;0·96] |
| ≥9 lung zones | 23/541 (4%) | 6/102 (6%) | 17/439 (4%) | 0·06 [0·03;0·12] | 0·96 [0·94;0·98] |
| ≥10 lung zones | 18/541 (3%) | 4/102 (4%) | 14/439 (3%) | 0·04 [0·02;0·1] | 0·97 [0·95;0·98] |
| ≥11 lung zones | 15/541 (3%) | 3/102 (3%) | 12/439 (3%) | 0·03 [0·01;0·08] | 0·97 [0·95;0·98] |
| ≥12 lung zones | 12/541 (2%) | 1/102 (1%) | 11/439 (3%) | 0·01 [0;0·05] | 0·97 [0·96;0·99] |
| ≥13 lung zones | 8/541 (1%) | 1/102 (1%) | 7/439 (2%) | 0·01 [0;0·05] | 0·98 [0·97;0·99] |
| ≥14 lung zones | 5/541 (1%) | 0/102 (0%) | 5/439 (1%) | 0 [0;0·04] | 0·99 [0·97;1] |
| Other ultrasound targets | | | | | |
| Internal mammary lymph nodes (IMNs) ≥0·5cm present | 34/541 (6%) | 16/102 (16%) | 18/439 (4%) | 0·16 [0·1;0·24] | 0·96 [0·94;0·97] |
| maximum size of IMNs | 10 [8;12] (N=34) | 11 [8;13] (N=16) | 10 [8;12] (N=18) | NA | NA |
| side of IMNs | Unilateral: 26/34 (76)  bilateral: 8/34 (24) | Unilateral: 11/16 (69)  bilateral: 5/16 (31) | Unilateral: 15/18 (83)  bilateral: 3/18 (17) | NA | NA |
| Additional lymph node pathology | Necrosis: 4/34 (12)  bulking: 4/34 (12) | Necrosis: 2/16 (13)  bulking: 3/16 (19) | necrosis: 2/18 (11)  bulking: 1 (6) | NA | NA |
| Pleural Nodules | 5/541 (1%) | 1/102 (1%) | 4/439 (1%) | NA | NA |
| Pleural laminar thickening | 44/541 (8%) | 14/102 (14%) | 30/439 (7%) | NA | NA |
| Pleural nodules or laminar thickening | 44/541 (8%) | 14/102 (14%) | 30/439 (7%) | NA | NA |
| diameter of pleural nodule (mm) | 5 [3;5] (N=5) | 5 [5;5] (N=1) | 4 [3;6] (N=4) | NA | NA |
| laminar pleural thickening, maximum thickness (mm) | 7 [6;10] (N=44) | 8 [6;9] (N=14) | 7 [5;10] (N=30) | NA | NA |
| Intestinal thickening in the right lower quadrant >4mm | 15/530 (3%) | 9/102 (9%) | 6/428 (1%) | 0·09 [0·05;0·16] | 0·99 [0·97;0·99] |
| maximum size of intestinal thickening | 5 [4;6] (N=15) | 5 [4;6] (N=9) | 5 [4;6] (N=6) | NA | NA |
| Any peritoneal thickening * | 14/541 (3%) | 11/102 (11%) | 3/439 (1%) | 0·11 [0·06;0·18] | 0·99 [0·98;1] |
| Mediastinal lymph nodes seen from suprasternal view ** | 2/540 (0%) | 0/102 (0%) | 2/438 (0%) | NA | NA |
| Peripheral lymph nodes present (only if clinical suspicion) | 3/539 (1%) | 1/102 (1%) | 2/437 (0%) | NA | NA |
| **Legend**  denominators provided for all individuals with available data for each line·  * This affected the omentum in most cases (10/11) with additional hypoechoic nodules in 8/10  ** parasternal only negative  IQR, interquartile range;  n, number;  MRS, microbiological reference standard;  eMRS, extended MRS;  CRS, composite reference standard;  TB, tuberculoisis;  FASH, focused assessment with sonography for HIV-associated tuberculosis;  SPC, subpleural consolidation;  LUS, lung ultrasound;  IMN, internal mammars lymph node  NA, not applicable | | | | | |

| **Table 4 - POCUS – stratified by HIV and diabetes** | | | | | | | | | | | | |
| --- | --- | --- | --- | --- | --- | --- | --- | --- | --- | --- | --- | --- |
|  | DM-, all  (n=391) | DM-, CRS+ (n=71) | DM-, unlikely (n=320) | DM+, all (n=121) | DM+, CRS+ (n=23) | DM+, unlikely (n=98) | HIV-, all (n=517) | HIV-, CRS+ (n=100) | HIV-, unlikely (n=417) | HIV+, all (n=6) | HIV+, CRS+ (n=1) | HIV+, unlikely (n=5) |
| Diagnostics | | | | | | | | | | | | |
| CXR suggestive of active TB | 54/383 (14%) | 24/69 (35%) | 30/314 (10%) | 24/121 (20%) | 9/23 (39%) | 15/98 (15%) | 78/509 (15%) | 33/98 (34%) | 45/411 (11%) | 1/6 (17%) | 1/1 (100%) | 0/5 (0%) |
| CXR suggestive or consistent with active TB | 180/383 (47%) | 56/69 (81%) | 124/314 (39%) | 74/121 (61%) | 20/23 (87%) | 54/98 (55%) | 255/509 (50%) | 79/98 (81%) | 176/411 (43%) | 3/6 (50%) | 1/1 (100%) | 2/5 (40%) |
| Positive TB-PCR on sputum or BAL | 32/391 (8%) | 32/71 (45%) | 0/320 (0%) | 14/121 (12%) | 14/23 (61%) | 0/98 (0%) | 46/517 (9%) | 46/100 (46%) | 0/417 (0%) | 1/6 (17%) | 1/1 (100%) | 0/5 (0%) |
| Positive TB-culture on sputum or BAL | 28/391 (7%) | 28/71 (39%) | 0/320 (0%) | 10/121 (8%) | 10/23 (43%) | 0/98 (0%) | 37/517 (7%) | 37/100 (37%) | 0/417 (0%) | 1/6 (17%) | 1/1 (100%) | 0/5 (0%) |
| Positive TB-PCR on non-sputum/BAL sample | 13/391 (3%) | 13/71 (18%) | 0/320 (0%) | 4/121 (3%) | 4/23 (17%) | 0/98 (0%) | 19/517 (4%) | 19/100 (19%) | 0/417 (0%) | 0/6 (0%) | 0/1 (0%) | 0/5 (0%) |
| Positive TB-culture on non-sputum/BAL sample | 8/391 (2%) | 8/71 (11%) | 0/320 (0%) | 3/121 (2%) | 3/23 (13%) | 0/98 (0%) | 11/517 (2%) | 11/100 (11%) | 0/417 (0%) | 0/6 (0%) | 0/1 (0%) | 0/5 (0%) |
| PTB only  EPTB only  PTB+EPTB | 34/71 (48)  14/71 (20)  23/71 (32) | | | 14/23 (61)  1/23 (4)  8/23 (35) | | | 50/100 (50)  16/100 (16)  34/100 (34 | | | 1/1 (100) PTB | | |
| FASH HEADLINE | | | | | | | | | | | | |
| FASH (original) positive | 129/391 (33%) | 39/71 (55%) | 90/320 (28%) | 45/121 (37%) | 10/23 (43%) | 35/98 (36%) | 180/517 (35%) | 51/100 (51%) | 129/417 (31%) | 1/6 (17%) | 1/1 (100%) | 0/5 (0%) |
| Pleural effusion present, any | 121/391 (31%) | 34/71 (48%) | 87/320 (27%) | 44/121 (36%) | 10/23 (43%) | 34/98 (35%) | 170/517 (33%) | 46/100 (46%) | 124/417 (30%) | 1/6 (17%) | 1/1 (100%) | 0/5 (0%) |
| Pericardial effusion ≥10mm | 5/391 (1%) | 2/71 (3%) | 3/320 (1%) | 2/121 (2%) | 0/23 (0%) | 2/98 (2%) | 8/517 (2%) | 2/100 (2%) | 6/417 (1%) | 0/6 (0%) | 0/1 (0%) | 0/5 (0%) |
| Hypoechoic spleen lesions <1·5cm present | 5/391 (1%) | 3/71 (4%) | 2/320 (1%) | 1/121 (1%) | 1/23 (4%) | 0/98 (0%) | 7/517 (1%) | 4/100 (4%) | 3/417 (1%) | 0/6 (0%) | 0/1 (0%) | 0/5 (0%) |
| Hypoechoic liver lesions | 4/391 (1%) | 0/71 (0%) | 4/320 (1%) | 1/121 (1%) | 0/23 (0%) | 1/98 (1%) | 5/517 (1%) | 0/100 (0%) | 5/417 (1%) | 0/6 (0%) | 0/1 (0%) | 0/5 (0%) |
| Abdominal lymph nodes ≥1·5cm present | 9/391 (2%) | 4/71 (6%) | 5/320 (2%) | 5/121 (4%) | 3/23 (13%) | 2/98 (2%) | 15/517 (3%) | 7/100 (7%) | 8/417 (2%) | 0/6 (0%) | 0/1 (0%) | 0/5 (0%) |
| Ascites present | 18/391 (5%) | 6/71 (8%) | 12/320 (4%) | 9/121 (7%) | 5/23 (22%) | 4/98 (4%) | 29/517 (6%) | 14/100 (14%) | 15/417 (4%) | 0/6 (0%) | 0/1 (0%) | 0/5 (0%) |
| LUNG ULTRASOUND | | | | | | | | | | | | |
| Subpleural consolidations (SPC) <1cm present | 333/391 (85%) | 67/71 (94%) | 266/320 (83%) | 108/121 (89%) | 22/23 (96%) | 86/98 (88%) | 443/517 (86%) | 93/100 (93%) | 350/417 (84%) | 5/6 (83%) | 1/1 (100%) | 4/5 (80%) |
| Subpleural consolidations ≥1cm present | 195/391 (50%) | 52/71 (73%) | 143/320 (45%) | 65/121 (54%) | 17/23 (74%) | 48/98 (49%) | 260/517 (50%) | 71/100 (71%) | 189/417 (45%) | 4/6 (67%) | 1/1 (100%) | 3/5 (60%) |
| any subpleural consolidation present, regardless of size? | 340/391 (87%) | 70/71 (99%) | 270/320 (84%) | 112/121 (93%) | 23/23 (100%) | 89/98 (91%) | 455/517 (88%) | 98/100 (98%) | 357/417 (86%) | 5/6 (83%) | 1/1 (100%) | 4/5 (80%) |
| miliary pattern present | 47/391 (12%) | 11/71 (15%) | 36/320 (11%) | 13/121 (11%) | 1/23 (4%) | 12/98 (12%) | 59/517 (11%) | 12/100 (12%) | 47/417 (11%) | 2/6 (33%) | 1/1 (100%) | 1/5 (20%) |
| B-lines (>2) in at least one lung zone | 182/391 (47%) | 40/71 (56%) | 142/320 (44%) | 49/121 (40%) | 8/23 (35%) | 41/98 (42%) | 235/517 (45%) | 52/100 (52%) | 183/417 (44%) | 5/6 (83%) | 1/1 (100%) | 4/5 (80%) |
| Other ultrasound targets | | | | | | | | | | | | |
| Internal mammary lymph nodes (IMNs) ≥0·5cm present | 28/391 (7%) | 13/71 (18%) | 15/320 (5%) | 5/121 (4%) | 2/23 (9%) | 3/98 (3%) | 34/517 (7%) | 16/100 (16%) | 18/417 (4%) | 0/6 (0%) | 0/1 (0%) | 0/5 (0%) |
| Pleural nodules or laminar thickening | 30/391 (8%) | 11/71 (15%) | 19/320 (6%) | 12/121 (10%) | 3/23 (13%) | 9/98 (9%) | 43/517 (8%) | 13/100 (13%) | 30/417 (7%) | 1/6 (17%) | 1/1 (100%) | 0/5 (0%) |
| Intestinal thickening in the right lower quadrant >4mm | 9/385 (2%) | 6/71 (8%) | 3/314 (1%) | 2/117 (2%) | 1/23 (4%) | 1/94 (1%) | 14/506 (3%) | 9/100 (9%) | 5/406 (1%) | 0/6 (0%) | 0/1 (0%) | 0/5 (0%) |
| Any peritoneal thickening | 6/391 (2%) | 4/71 (6%) | 2/320 (1%) | 6/121 (5%) | 5/23 (22%) | 1/98 (1%) | 14/517 (3%) | 11/100 (11%) | 3/417 (1%) | 0/6 (0%) | 0/1 (0%) | 0/5 (0%) |
| Peripheral lymph nodes present (only if clinical suspicion) | 2/390 (1%) | 0/71 (0%) | 2/319 (1%) | 1/120 (1%) | 1/23 (4%) | 0/97 (0%) | 3/515 (1%) | 1/100 (1%) | 2/415 (0%) | 0/6 (0%) | 0/1 (0%) | 0/5 (0%) |
| **Legend**  denominators provided for all individuals with available data for each line·  DM, diabetes mellitus;  n, number;  HIV, human immunodeficiency virus;  CRS, composite reference standard;  CXR, chest x-ray;  TB, tuberculosis;  PCR, polymerase chain reaction;  BAL, broncho-alveolar lavage;  EPTB, extra-pulmonary tuberculosis;  PTB, pulmonary tuberculosis;  FASH, focused assessment with sonography for HIV-associated tuberculosis;  SPC, subpleural consolidations;  IMN, internal mammary lymph nodes | | | | | | | | | | | | |

| **Supplement Table 5 PP data for cohort characteristics, test results and index test** | | | | | |
| --- | --- | --- | --- | --- | --- |
|  | **ALL** | **MRS** | **eMRS** | **CRS** | **Unlikely TB** |
| N | 489 | 54 | 67 | 92 | 397 |
| PATIENT CHARACTERISTICS | | | | | |
| Age in years | 48 [35;60] (N=489) | 46 [30;57] (N=54) | 46 [30;57] (N=67) | 42 [29;55] (N=92) | 49 [36;60] (N=397) |
| Gender male | 324/489 (66%) | 36/54 (67%) | 43/67 (64%) | 60/92 (65%) | 264/397 (66%) |
| Body mass index (kg/m^2) | 21·7 [18·6;25·1] (N=489) | 19·1 [17·3;22·4] (N=54) | 19 [17;22·6] (N=67) | 19·3 [17·5;23·4] (N=92) | 22·1 [19·2;25·4] (N=397) |
| History of tobacco use | 95/489 (19%) | 10/54 (19%) | 10/67 (15%) | 16/92 (17%) | 79/397 (20%) |
| Final diabetes status | 113/462 (24%) | 16/51 (31%) | 18/62 (29%) | 22/84 (26%) | 91/378 (24%) |
| Final HIV status | 6/472 (1%) | 1/54 (2%) | 1/67 (1%) | 1/92 (1%) | 5/380 (1%) |
| History of previous TB disease | 87/488 (18%) | 6/54 (11%) | 8/67 (12%) | 9/92 (10%) | 78/396 (20%) |
| History of current TB-contact | 5/483 (1%) | 1/54 (2%) | 1/67 (1%) | 3/92 (3%) | 2/391 (1%) |
| CURRENT SYMPTOMS | | | | | |
| Cough | 440/489 (90%) | 52/54 (96%) | 62/67 (93%) | 82/92 (89%) | 358/397 (90%) |
| Hemoptysis | 109/489 (22%) | 9/54 (17%) | 12/67 (18%) | 15/92 (16%) | 94/397 (24%) |
| Night sweats | 46/489 (9%) | 10/54 (19%) | 12/67 (18%) | 16/92 (17%) | 30/397 (8%) |
| Fever | 207/489 (42%) | 39/54 (72%) | 48/67 (72%) | 63/92 (68%) | 144/397 (36%) |
| Weight loss | 259/489 (53%) | 39/54 (72%) | 48/67 (72%) | 67/92 (73%) | 192/397 (48%) |
| Fatigue | 316/489 (65%) | 38/54 (70%) | 49/67 (73%) | 65/92 (71%) | 251/397 (63%) |
| Loss of appetite | 220/489 (45%) | 33/54 (61%) | 42/67 (63%) | 60/92 (65%) | 160/397 (40%) |
| Abdominal pain or distension | 82/489 (17%) | 8/54 (15%) | 11/67 (16%) | 22/92 (24%) | 60/397 (15%) |
| Peripheral lymph node swelling | 6/489 (1%) | 1/54 (2%) | 3/67 (4%) | 4/92 (4%) | 2/397 (1%) |
| DIAGNOSTICS | | | | | |
| HbA1c (%) | 6 [5;6] (N=460) | 6 [5;6] (N=50) | 6 [5;6] (N=61) | 6 [5;6] (N=83) | 6 [5;6] (N=377) |
| C-reactive protein (mg/l) | 6 [3;23] (N=437) | 30 [6;58] (N=48) | 30 [7;60] (N=59) | 26 [8;59] (N=83) | 4 [3;16] (N=354) |
| CRP > 5 | 232/437 (53%) | 37/48 (77%) | 47/59 (80%) | 68/83 (82%) | 164/354 (46%) |
| CXR suggestive of active TB | 67/480 (14%) | 21/52 (40%) | 24/65 (37%) | 28/90 (31%) | 39/390 (10%) |
| CXR suggestive or consistent with active TB | 231/480 (48%) | 44/52 (85%) | 54/65 (83%) | 72/90 (80%) | 159/390 (41%) |
| CXR suggestive of post-TB (with or without signs of active TB) | 129/476 (27%) | 18/51 (35%) | 21/63 (33%) | 24/87 (28%) | 105/389 (27%) |
| TB-PCR on urine positive | 5/489 (1%) | 5/54 (9%) | 5/67 (7%) | 5/92 (5%) | 0/397 (0%) |
| Number of sputa investigated for TB | 1: 17; 2: 353; 3: 112; 4: 7 | 1: 3; 2: 33; 3: 14; 4: 4 | 1: 4; 2: 41; 3: 18; 4: 4 | 1: 4; 2: 55; 3: 29; 4: 4 | 1: 13; 2: 298; 3: 83; 4: 3 |
| Number of patients for whom bronchoalveolar lavage was investigated for TB | 129/489 (26%) | 20/54 (37%) | 22/67 (33%) | 26/92 (28%) | 103/397 (26%) |
| Number of patients for whom extra-pulmonary samples were investigated for TB | 178/489 (36%) | 21/54 (39%) | 34/67 (51%) | 57/92 (62%) | 121/397 (30%) |
| Positive TB-PCR or TB-cult on sputum or BAL | 47/489 (10%) | 47/54 (87%) | 47/67 (70%) | 47/92 (51%) | 0/397 (0%) |
| Positive TB-PCR or TB-Cult on EPTB sample | 20/489 (4%) | 7/54 (13%) | 20/67 (30%) | 20/92 (22%) | 0/397 (0%) |
| PTB, EPTB or concurrent PTB+EPTB? | 1: 48; 2: 14; 3: 30 | 1: 43; 2: 1; 3: 10 | 1: 46; 2: 3; 3: 18 | 1: 48; 2: 14; 3: 30 |  |
| FASH HEADLINE | | | | | |
| FASH (original) positive | 156/489 (32%) | 18/54 (33%) | 24/67 (36%) | 46/92 (50%) | 110/397 (28%) |
| FASH (original with ascites) positive | 160/489 (33%) | 18/54 (33%) | 24/67 (36%) | 47/92 (51%) | 113/397 (28%) |
| FASH (original with 4mm cut-off for pericardial effusion) | 170/489 (35%) | 19/54 (35%) | 25/67 (37%) | 48/92 (52%) | 122/397 (31%) |
| FASH (original with 200ml cut-off for pleural effusion) | 136/489 (28%) | 16/54 (30%) | 22/67 (33%) | 44/92 (48%) | 92/397 (23%) |
| FASH (original with 300ml cut-off for pleural effusion) | 116/489 (24%) | 12/54 (22%) | 17/67 (25%) | 39/92 (42%) | 77/397 (19%) |
| FASH (original with 400ml cut-off for pleural effusion) | 102/489 (21%) | 12/54 (22%) | 17/67 (25%) | 37/92 (40%) | 65/397 (16%) |
| FASH (original with 600ml cut-off for pleural effusion) | 92/489 (19%) | 9/54 (17%) | 14/67 (21%) | 33/92 (36%) | 59/397 (15%) |
| FASH (original with 800ml cut-off for pleural effusion) | 85/489 (17%) | 9/54 (17%) | 14/67 (21%) | 31/92 (34%) | 54/397 (14%) |
| FASH (original with 1000ml cut-off for pleural effusion) | 75/489 (15%) | 7/54 (13%) | 12/67 (18%) | 28/92 (30%) | 47/397 (12%) |
| Pleural effusion present, any | 146/489 (30%) | 17/54 (31%) | 22/67 (33%) | 41/92 (45%) | 105/397 (26%) |
| Pericardial effusion >=4mm | 34/489 (7%) | 3/54 (6%) | 4/67 (6%) | 7/92 (8%) | 27/397 (7%) |
| Pericardial effusion >=10mm | 7/489 (1%) | 0/54 (0%) | 1/67 (1%) | 2/92 (2%) | 5/397 (1%) |
| Hypoechoic spleen lesions <1·5cm present | 7/489 (1%) | 1/54 (2%) | 2/67 (3%) | 4/92 (4%) | 3/397 (1%) |
| Larger splenic lesions present (>=1·5cm) suggestive of macroabscess | 3/488 (1%) | 0/54 (0%) | 0/67 (0%) | 2/92 (2%) | 1/396 (0%) |
| Hyperechoic spleen lesions present with or without calcification | 23/488 (5%) | 4/54 (7%) | 5/67 (7%) | 5/92 (5%) | 18/396 (5%) |
| Size of spleen | 10 [9;11] (N=479) | 10 [9;11] (N=54) | 10 [9;11] (N=67) | 10 [9;11] (N=92) | 10 [9;11] (N=387) |
| Hypoechoic liver lesions | 5/489 (1%) | 0/54 (0%) | 0/67 (0%) | 0/92 (0%) | 5/397 (1%) |
| Abdominal lymph nodes >=1·5cm present | 14/489 (3%) | 3/54 (6%) | 4/67 (6%) | 7/92 (8%) | 7/397 (2%) |
| Ascites present | 25/489 (5%) | 4/54 (7%) | 7/67 (10%) | 13/92 (14%) | 12/397 (3%) |
| LUNG ULTRASOUND | | | | | |
| <1cm consolidations | | | | | |
| Subpleural consolidations (SPC) <1cm present | 416/489 (85%) | 50/54 (93%) | 63/67 (94%) | 86/92 (93%) | 330/397 (83%) |
| SPCs with regular round/oval shape, hypoechoic echogenicity and posterior enhancement | 11/489 (2%) | 0/54 (0%) | 0/67 (0%) | 1/92 (1%) | 10/397 (3%) |
| Number of subpleural consolidations <1cm | 1: 60; 2: 206; 3: 150 | 1: 2; 2: 26; 3: 22 | 1: 4; 2: 32; 3: 27 | 1: 9; 2: 43; 3: 34 | 1: 51; 2: 163; 3: 116 |
| Number of lung zones with subpleural consolidations <1cm | 4 [2;6] (N=416) | 4 [3;6] (N=50) | 4 [3;6] (N=63) | 4 [3;5] (N=86) | 4 [2;7] (N=330) |
| >5 SPCs present | 150/489 (31%) | 22/54 (41%) | 27/67 (40%) | 34/92 (37%) | 116/397 (29%) |
| SPCs with >=5mm present | 320/489 (65%) | 42/54 (78%) | 53/67 (79%) | 70/92 (76%) | 250/397 (63%) |
| >5 SPCs with at least one >=5mm | 131/489 (27%) | 20/54 (37%) | 24/67 (36%) | 30/92 (33%) | 101/397 (25%) |
| >=1cm consolidations | | | | | |
| Subpleural consolidations >=1cm present | 240/489 (49%) | 38/54 (70%) | 45/67 (67%) | 65/92 (71%) | 175/397 (44%) |
| Number of lung zones with subpleural consolidations >=1cm | 2 [1;4] (N=240) | 3 [2;4] (N=38) | 2 [1;4] (N=45) | 3 [2;4] (N=65) | 2 [1;4] (N=175) |
| combinations of <1cm and >=1cm | | | | | |
| any subpleural consolidation present, regardless of size? | 426/489 (87%) | 53/54 (98%) | 66/67 (99%) | 90/92 (98%) | 336/397 (85%) |
| Location of consolidations | | | | | |
| any consolidation in the apical regions | 261/489 (53%) | 40/54 (74%) | 49/67 (73%) | 59/92 (64%) | 202/397 (51%) |
| any consolidations <1cm in the apical regions | 224/489 (46%) | 33/54 (61%) | 42/67 (63%) | 52/92 (57%) | 172/397 (43%) |
| any consolidations >=1cm in the apical regions | 73/489 (15%) | 14/54 (26%) | 16/67 (24%) | 20/92 (22%) | 53/397 (13%) |
| any consollidations >=1cm in the upper lung zones | 163/489 (33%) | 32/54 (59%) | 37/67 (55%) | 48/92 (52%) | 115/397 (29%) |
| Other LUS findings | | | | | |
| miliary pattern present | 56/489 (11%) | 8/54 (15%) | 9/67 (13%) | 12/92 (13%) | 44/397 (11%) |
| B-lines (>2) in at least one lung zone | 218/489 (45%) | 29/54 (54%) | 35/67 (52%) | 48/92 (52%) | 170/397 (43%) |
| LUS findings depending on the number of zones affected | | | | | |
| At least 1 lung zone with subpleural consolidation <1cm | 416/489 (85%) | 50/54 (93%) | 63/67 (94%) | 86/92 (93%) | 330/397 (83%) |
| At least 2 lung zones with subpleural consolidation <1cm | 352/489 (72%) | 48/54 (89%) | 58/67 (87%) | 76/92 (83%) | 276/397 (70%) |
| At least 3 lung zones with subpleural consolidation <1cm | 284/489 (58%) | 41/54 (76%) | 51/67 (76%) | 65/92 (71%) | 219/397 (55%) |
| At least 4 lung zones with subpleural consolidation <1cm | 219/489 (45%) | 27/54 (50%) | 35/67 (52%) | 45/92 (49%) | 174/397 (44%) |
| At least 5 lung zones with subpleural consolidation <1cm | 163/489 (33%) | 19/54 (35%) | 26/67 (39%) | 33/92 (36%) | 130/397 (33%) |
| At least 6 lung zones with subpleural consolidation <1cm | 129/489 (26%) | 14/54 (26%) | 19/67 (28%) | 20/92 (22%) | 109/397 (27%) |
| At least 7 lung zones with subpleural consolidation <1cm | 100/489 (20%) | 11/54 (20%) | 14/67 (21%) | 15/92 (16%) | 85/397 (21%) |
| At least 8 lung zones with subpleural consolidation <1cm | 72/489 (15%) | 11/54 (20%) | 13/67 (19%) | 14/92 (15%) | 58/397 (15%) |
| At least 9 lung zones with subpleural consolidation <1cm | 47/489 (10%) | 8/54 (15%) | 10/67 (15%) | 11/92 (12%) | 36/397 (9%) |
| At least 10 lung zones with subpleural consolidation <1cm | 32/489 (7%) | 7/54 (13%) | 8/67 (12%) | 9/92 (10%) | 23/397 (6%) |
| At least 11 lung zones with subpleural consolidation <1cm | 23/489 (5%) | 5/54 (9%) | 5/67 (7%) | 6/92 (7%) | 17/397 (4%) |
| At least 12 lung zones with subpleural consolidation <1cm | 15/489 (3%) | 3/54 (6%) | 3/67 (4%) | 4/92 (4%) | 11/397 (3%) |
| At least 13 lung zones with subpleural consolidation <1cm | 9/489 (2%) | 1/54 (2%) | 1/67 (1%) | 2/92 (2%) | 7/397 (2%) |
| At least 14 lung zones with subpleural consolidation <1cm | 6/489 (1%) | 0/54 (0%) | 0/67 (0%) | 1/92 (1%) | 5/397 (1%) |
| At least 1 lung zone with subpleural consolidation >=1cm | 240/489 (49%) | 38/54 (70%) | 45/67 (67%) | 65/92 (71%) | 175/397 (44%) |
| At least 2 lung zones with subpleural consolidation >=1cm | 166/489 (34%) | 29/54 (54%) | 33/67 (49%) | 50/92 (54%) | 116/397 (29%) |
| At least 3 lung zones with subpleural consolidation >=1cm | 108/489 (22%) | 20/54 (37%) | 22/67 (33%) | 36/92 (39%) | 72/397 (18%) |
| At least 4 lung zones with subpleural consolidation >=1cm | 73/489 (15%) | 15/54 (28%) | 17/67 (25%) | 25/92 (27%) | 48/397 (12%) |
| At least 5 lung zones with subpleural consolidation >=1cm | 44/489 (9%) | 9/54 (17%) | 9/67 (13%) | 11/92 (12%) | 33/397 (8%) |
| At least 6 lung zones with subpleural consolidation >=1cm | 27/489 (6%) | 4/54 (7%) | 4/67 (6%) | 5/92 (5%) | 22/397 (6%) |
| At least 7 lung zones with subpleural consolidation >=1cm | 19/489 (4%) | 4/54 (7%) | 4/67 (6%) | 4/92 (4%) | 15/397 (4%) |
| At least 8 lung zones with subpleural consolidation >=1cm | 10/489 (2%) | 4/54 (7%) | 4/67 (6%) | 4/92 (4%) | 6/397 (2%) |
| At least 9 lung zones with subpleural consolidation >=1cm | 4/489 (1%) | 1/54 (2%) | 1/67 (1%) | 1/92 (1%) | 3/397 (1%) |
| At least 10 lung zones with subpleural consolidation >=1cm | 1/489 (0%) | 0/54 (0%) | 0/67 (0%) | 0/92 (0%) | 1/397 (0%) |
| At least 11 lung zones with subpleural consolidation >=1cm | 1/489 (0%) | 0/54 (0%) | 0/67 (0%) | 0/92 (0%) | 1/397 (0%) |
| At least 12 lung zones with subpleural consolidation >=1cm | 0/489 (0%) | 0/54 (0%) | 0/67 (0%) | 0/92 (0%) | 0/397 (0%) |
| At least 13 lung zones with subpleural consolidation >=1cm | 0/489 (0%) | 0/54 (0%) | 0/67 (0%) | 0/92 (0%) | 0/397 (0%) |
| At least 14 lung zones with subpleural consolidation >=1cm | 0/489 (0%) | 0/54 (0%) | 0/67 (0%) | 0/92 (0%) | 0/397 (0%) |
| At least 1 lung zone with B-lines | 218/489 (45%) | 29/54 (54%) | 35/67 (52%) | 48/92 (52%) | 170/397 (43%) |
| At least 2 lung zones with B-lines | 156/489 (32%) | 21/54 (39%) | 26/67 (39%) | 34/92 (37%) | 122/397 (31%) |
| At least 3 lung zones with B-lines | 107/489 (22%) | 16/54 (30%) | 18/67 (27%) | 23/92 (25%) | 84/397 (21%) |
| At least 4 lung zones with B-lines | 81/489 (17%) | 12/54 (22%) | 14/67 (21%) | 19/92 (21%) | 62/397 (16%) |
| At least 5 lung zones with B-lines | 63/489 (13%) | 11/54 (20%) | 11/67 (16%) | 14/92 (15%) | 49/397 (12%) |
| At least 6 lung zones with B-lines | 45/489 (9%) | 9/54 (17%) | 9/67 (13%) | 10/92 (11%) | 35/397 (9%) |
| At least 7 lung zones with B-lines | 37/489 (8%) | 7/54 (13%) | 7/67 (10%) | 8/92 (9%) | 29/397 (7%) |
| At least 8 lung zones with B-lines | 29/489 (6%) | 7/54 (13%) | 7/67 (10%) | 8/92 (9%) | 21/397 (5%) |
| At least 9 lung zones with B-lines | 21/489 (4%) | 5/54 (9%) | 5/67 (7%) | 6/92 (7%) | 15/397 (4%) |
| At least 10 lung zones with B-lines | 16/489 (3%) | 4/54 (7%) | 4/67 (6%) | 4/92 (4%) | 12/397 (3%) |
| At least 11 lung zones with B-lines | 13/489 (3%) | 3/54 (6%) | 3/67 (4%) | 3/92 (3%) | 10/397 (3%) |
| At least 12 lung zones with B-lines | 10/489 (2%) | 1/54 (2%) | 1/67 (1%) | 1/92 (1%) | 9/397 (2%) |
| At least 13 lung zones with B-lines | 8/489 (2%) | 1/54 (2%) | 1/67 (1%) | 1/92 (1%) | 7/397 (2%) |
| At least 14 lung zones with B-lines | 5/489 (1%) | 0/54 (0%) | 0/67 (0%) | 0/92 (0%) | 5/397 (1%) |
| OTHER US TARGETS | | | | | |
| Internal mammary lymph nodes (IMNs) >=0·5cm present | 29/489 (6%) | 5/54 (9%) | 9/67 (13%) | 14/92 (15%) | 15/397 (4%) |
| max. size of IMNs | 10 [8;12] (N=29) | 9 [8;11] (N=5) | 11 [8;12] (N=9) | 11 [10;13] (N=14) | 10 [8;12] (N=15) |
| side of IMNs | 1: 14; 2: 8; 3: 7 | 1: 2; 2: 1; 3: 2 | 1: 4; 2: 2; 3: 3 | 1: 5; 2: 4; 3: 5 | 1: 9; 2: 4; 3: 2 |
| Pleural laminar thickening | 39/489 (8%) | 6/54 (11%) | 7/67 (10%) | 12/92 (13%) | 27/397 (7%) |
| Intestinal thickening in the right lower quadrant >4mm | 15/478 (3%) | 3/54 (6%) | 5/67 (7%) | 9/92 (10%) | 6/386 (2%) |
| Any peritoneal thickening (parietal, visceral, omental) | 13/489 (3%) | 4/54 (7%) | 6/67 (9%) | 10/92 (11%) | 3/397 (1%) |
| Mediastinal lymph nodes seen from suprasternal view [parasternal only negative] | 2/488 (0%) | 0/54 (0%) | 0/67 (0%) | 0/92 (0%) | 2/396 (1%) |
| Peripheral lymph nodes present (only if clinical suspicion) | 3/487 (1%) | 1/54 (2%) | 1/67 (1%) | 1/92 (1%) | 2/395 (1%) |
| **Legend**  denominators provided for all individuals with available data for each line.  IQR, interquartile range;  n, number;  MRS, microbiological reference standard;  eMRS, extended MRS;  CRS, composite reference standard;  TB, tuberculoisis;  WHO, World Health Organization;  HIV, human immunodeficiency virus;  ART, anti-retroviral therapy;  CXR, chest x-ray;  PCR, polymerase chain reaction;  BAL, broncho-alveolar lavage;  EPTB, extra-pulmonary tuberculosis  FASH, focused assessment with sonography for HIV-associated tuberculosis;  SPC, subpleural consolidation;  LUS, lung ultrasound;  IMN, internal mammars lymph node  NA, not applicable | | | | | |

Interrater analysis

Interobserver agreement calculations (see methods) yielded an overall Cohen’s kappa of 0·71, and more specifically for rater 1 and 2: 0·65, raters 1 and 3: 0·79, and raters 2 and 3: 0·72. From the generalized linear mixed we inferred the probability of agreement to be 98·8%, which strongly supports coherence in rater decisions.

Predictive modelling analysis

Variables in factor analysis: participant variables (age, gender, diabetes, HIV, C-reactive protein, history of fever, history of weight loss), ultrasound variables (pleural fluid volume estimate, pericardial effusion, hypoechoic spleen lesions, hyperechoic spleen lesions, spleen size, hypoechoic liver lesions, abdominal lymphadenopathy, SPC_≥1cm_, SPC_≥1cm_ in the apical region, SPC_<1cm_, SPC_<1cm_ in the apical regions, miliary pattern, internal mammary lymph nodes, pleural thickening, peritoneal thickening)

Lasso variables: participants variables (age, history of fever, history of weight loss, C-reactive protein), ultrasound variables (pleural fluid volume estimate, spleen size, SPC_≥1cm_, SPC_<1cm_, SPC_<1cm_ in the apical regions, peritoneal thickening)

The loading matrix and the eigenvalues from the factor analysis is as follows:

| **Variable** | **Factor 1** | **Factor 2** | **Factor 3** | **Factor 4** | **Factor 5** | **Factor 6** | **Factor 7** | **Factor 8** | **Factor 9** |
| --- | --- | --- | --- | --- | --- | --- | --- | --- | --- |
| Age | -0,0495 | 0,0985 | 0,1252 | 0,0158 | 0,0689 | -0,0374 | 0,0895 | -0,2296 | 0,6162 |
| Sex | -0,0502 | -0,119 | 0,0139 | 0,0903 | -0,0349 | 0,0213 | 0,0772 | -0,0021 | -0,0895 |
| diabetes_final | 0,0471 | -0,0266 | -0,0921 | -0,0387 | -0,0605 | 0,0414 | 0,0452 | 0,0882 | 0,5004 |
| hiv_final | -0,0023 | -0,0189 | 0,0488 | -0,0439 | -0,0019 | -0,0054 | 0,0017 | -0,0231 | -0,0753 |
| cxr_tb_diag | -0,0469 | -0,0531 | 0,1389 | -0,0032 | -0,0189 | -0,0656 | -0,1286 | 0,5008 | 0,061 |
| FASH_ORIG | 0,1586 | 0,2903 | 0,0743 | 0,8292 | -0,0133 | 0,0921 | -0,0625 | -0,0223 | -0,0094 |
| PLEFF | 0,2407 | 0,2727 | 0,0400 | 0,9005 | 0,0049 | -0,0987 | -0,1091 | -0,0339 | 0,0232 |
| PLEFF.QUANT | -0,0480 | 0,9805 | 0,0746 | 0,286 | -0,1043 | -0,1669 | -0,0168 | -0,06 | -0,0266 |
| PERI | -0,0424 | 0,0864 | 0,0064 | 0,0255 | -0,0349 | 0,0883 | 0,5194 | -0,054 | 0,2242 |
| PERI_1CM | -0,0321 | 0,0225 | 0,039 | -0,1846 | 0,0031 | -0,0272 | 0,947 | 0,1719 | -0,0337 |
| spl_hypo | -0,0652 | -0,0623 | 0,0468 | -0,0699 | 0,0533 | 0,7182 | 0,0444 | -0,0347 | -0,0177 |
| spl_hyper | -0,025 | -0,0816 | 0,0952 | 0,0025 | -0,0517 | -0,0242 | -0,0077 | 0,0751 | 0,1298 |
| spl_size | 0,113 | 0,0725 | -0,1218 | -0,0045 | -0,1353 | 0,1285 | -0,0313 | 0,224 | -0,0883 |
| LIV | -0,1195 | 0,0129 | 0,0043 | 0,09 | -0,012 | 0,2095 | -0,0235 | 0,0429 | -0,0441 |
| ABDOM_LN | 0,2997 | 0,0192 | 0,0557 | -0,0168 | -0,0668 | 0,522 | -0,0566 | -0,022 | 0,017 |
| PERI_FLUID | 0,567 | 0,0358 | 0,0029 | 0,0744 | 0,0323 | 0,2113 | 0,1172 | -0,1241 | -0,008 |
| SUN_size5or_CONSOL | 0,0401 | -0,0295 | 0,3301 | 0,2116 | 0,2637 | 0,0724 | -0,0432 | 0,1197 | 0,1215 |
| sun_apex | 0,0688 | 0,0738 | 0,6227 | 0,0858 | -0,1805 | -0,0572 | 0,0239 | -0,0499 | -0,001 |
| MILIARY | 0,0399 | 0,0213 | 0,3723 | -0,0814 | 0,1554 | 0,0594 | 0,0146 | 0,0741 | -0,0891 |
| mamm_ln | 0,1973 | 0,0924 | 0,0509 | 0,0866 | 0,0776 | -0,1816 | 0,153 | 0,2009 | -0,1609 |
| PLNODLAM | -0,165 | 0,7359 | 0,0552 | 0,0346 | -0,0468 | -0,1197 | 0,0422 | -0,1025 | 0,052 |
| PERITANY | 1,062 | -0,1741 | 0,0435 | 0,1331 | -0,0588 | -0,0719 | -0,0113 | -0,0302 | 0,0143 |
| PERITTHICK_OMENT | 1,099 | -0,2214 | 0,0261 | 0,1504 | -0,0357 | -0,0263 | -0,1262 | -0,046 | 0,0139 |
| PERIPH | 0,1538 | -0,1249 | 0,0093 | 0,01 | 0,0651 | 0,4836 | 0,0432 | -0,1226 | 0,1031 |
| crp | 0,0054 | -0,0442 | -0,0844 | -0,0222 | 0,0982 | -0,1085 | 0,1912 | 0,5632 | 0,1547 |
| sx_fever | -0,0651 | -0,0763 | -0,0565 | 0,0432 | -0,0269 | -0,0178 | 0,0248 | 0,4248 | -0,0726 |
| sx_weight | 0,0321 | 0,0504 | 0,0525 | -0,1053 | -0,015 | 0,1008 | 0,0291 | 0,2377 | 0,0388 |
| sun | -0,0711 | 0,0914 | 1,053 | -0,0449 | -0,0691 | 0,1659 | 0,013 | -0,0277 | -0,1054 |
| FASH_strict600 | -0,1333 | 0,8182 | 0,0628 | 0,1434 | -0,0676 | 0,2496 | 0,1088 | -0,0083 | -0,0244 |
| consol_apexYN | -0,0289 | -0,1637 | -0,1634 | -0,065 | 0,9241 | 0,025 | 0,0064 | -0,1123 | -0,0407 |
| consolAtLeast3 | -0,0806 | 0,1823 | -0,0345 | 0,0135 | 0,5403 | 0,0193 | -0,034 | 0,2644 | -0,1055 |
| consolYN | -0,0342 | 0,0174 | 0,0667 | 0,1762 | 0,5829 | 0,0585 | -0,0604 | 0,2108 | 0,053 |
| *Eigenvalue:* | *5.45594998* | *3.4667042* | *2.64697761* | *2.30000521* | *2.20417654* | *1.95531362* | *1.74091252* | *1.52212533* | *1.28173286* |

|  | **Section & Topic** | **No** | **Item** | **Reported on page #** |
| --- | --- | --- | --- | --- |
|  | **TITLE OR ABSTRACT** |  |  |  |
|  |  | **1** | Identification as a study of diagnostic accuracy using at least one measure of accuracy  (such as sensitivity, specificity, predictive values, or AUC) | 1 |
|  | **ABSTRACT** |  |  |  |
|  |  | **2** | Structured summary of study design, methods, results, and conclusions  (for specific guidance, see STARD for Abstracts) | 2 |
|  | **INTRODUCTION** |  |  |  |
|  |  | **3** | Scientific and clinical background, including the intended use and clinical role of the index test | 4 |
|  |  | **4** | Study objectives and hypotheses | 4 |
|  | **METHODS** |  |  |  |
|  | *Study design* | **5** | Whether data collection was planned before the index test and reference standard  were performed (prospective study) or after (retrospective study) | 5 |
|  | *Participants* | **6** | Eligibility criteria | 5 |
|  |  | **7** | On what basis potentially eligible participants were identified  (such as symptoms, results from previous tests, inclusion in registry) | 5 |
|  |  | **8** | Where and when potentially eligible participants were identified (setting, location and dates) | 5 |
|  |  | **9** | Whether participants formed a consecutive, random or convenience series | 5 |
|  | *Test methods* | **10a** | Index test, in sufficient detail to allow replication | 5-6 |
|  |  | **10b** | Reference standard, in sufficient detail to allow replication | 6 |
|  |  | **11** | Rationale for choosing the reference standard (if alternatives exist) | 6 |
|  |  | **12a** | Definition of and rationale for test positivity cut-offs or result categories  of the index test, distinguishing pre-specified from exploratory | 5 |
|  |  | **12b** | Definition of and rationale for test positivity cut-offs or result categories  of the reference standard, distinguishing pre-specified from exploratory | 6 |
|  |  | **13a** | Whether clinical information and reference standard results were available  to the performers/readers of the index test | 6 |
|  |  | **13b** | Whether clinical information and index test results were available  to the assessors of the reference standard | 6 |
|  | *Analysis* | **14** | Methods for estimating or comparing measures of diagnostic accuracy | 6-7 |
|  |  | **15** | How indeterminate index test or reference standard results were handled | 6 |
|  |  | **16** | How missing data on the index test and reference standard were handled | 7 |
|  |  | **17** | Any analyses of variability in diagnostic accuracy, distinguishing pre-specified from exploratory | 5-6, Table 1 |
|  |  | **18** | Intended sample size and how it was determined | 6 |
|  | **RESULTS** |  |  |  |
|  | *Participants* | **19** | Flow of participants, using a diagram | Fig. 2 |
|  |  | **20** | Baseline demographic and clinical characteristics of participants | Table 2 |
|  |  | **21a** | Distribution of severity of disease in those with the target condition | Table 2 |
|  |  | **21b** | Distribution of alternative diagnoses in those without the target condition | 10, Table 5 |
|  |  | **22** | Time interval and any clinical interventions between index test and reference standard | 6 |
|  | *Test results* | **23** | Cross tabulation of the index test results (or their distribution)  by the results of the reference standard | Table 3 |
|  |  | **24** | Estimates of diagnostic accuracy and their precision (such as 95% confidence intervals) | Table 3 |
|  |  | **25** | Any adverse events from performing the index test or the reference standard | 11 |
|  | **DISCUSSION** |  |  |  |
|  |  | **26** | Study limitations, including sources of potential bias, statistical uncertainty, and generalisability | 12 |
|  |  | **27** | Implications for practice, including the intended use and clinical role of the index test | 11, 12 |
|  | **OTHER INFORMATION** |  |  |  |
|  |  | **28** | Registration number and name of registry | 5 |
|  |  | **29** | Where the full study protocol can be accessed | Supplement |
|  |  | **30** | Sources of funding and other support; role of funders | 13 |
